# Distribution of capsule and O types in *Klebsiella pneumoniae* causing neonatal sepsis in Africa and South Asia: meta-analysis of genome-predicted serotype prevalence to inform potential vaccine coverage

**DOI:** 10.1101/2025.06.28.25330253

**Authors:** Thomas D Stanton, Shaun P Keegan, Jabir A Abdulahi, Anne V Amulele, Matthew Bates, Eva Heinz, Yogesh Hooda, Weiming Hu, Kajal Jain, Samiah Kanwar, Rindidzani Magobo, Courtney P Olwagen, John M Tembo, Tolbert Sonda, Jonathan Strysko, Caroline C Tigoi, Sameen Ahmad Amin, Kyle Bittinger, Jennifer Cornick, Ebenezer Foster-Nyarko, Wilson Gumbi, Aneeta Hotwani, Naveed Iqbal, Steven M Jones, Furqan Kabir, Waqasuddin Khan, Chileshe L Musyani, Carolyn M McGann, Varsha Mittal, Ahmed M Moustafa, Patrick Musicha, James CL Mwansa, Moreka L Ndumba, Erkison E Odih, Donwilliams O Omuoyo, Oliver Pearse, Laura T Phillips, Paul J Planet, Aniqa Abdul Rasool, Charlene MC Rodrigues, Kirsty Sands, Arif M Tanmoy, Erin Theiller, Allan M Zuza, Sulagna Basu, Grace J Chan, Kenneth C Iregbu, Jean-Baptiste Mazarati, Semaria Solomon Alemayehu, Timothy R Walsh, Rabaab Zahra, Angela Dramowski, Sombo Fwoloshi, Appiah-Korang Labi, Lola Madrid, Noah Obeng-Nkrumah, David Ojok, Boaz D Wadugu, Andrew C Whitelaw, Adhisivam Bethou, Anudita Bhargava, Atul Jindal, Ruchi N Nanavati, Priyanka S Prasad, Apurba Sastry, Joveria Q Farooqi, Najia Ghanchi, Fyezah Jehan, Erum Khan, Ramesh K Agarwal, Alexander M Aiken, James A Berkley, Susan E Coffin, Nicholas A Feasey, Nelesh P Govender, Davidson H Hamer, Shabir A Madhi, M Imran Nisar, Samir K Saha, Senjuti Saha, M Jeeva Sankar, Kelly L Wyres, Kathryn E Holt

## Abstract

**Background:** *Klebsiella pneumoniae* causes ∼20% of sepsis in neonates, with ∼40% crude mortality. A vaccine administered to pregnant women, protecting against ≥70% of *K. pneumoniae* infections, could avert ∼400,000 cases and ∼80,000 deaths annually, mostly in Africa and South Asia. Vaccine formulations targeting the capsular polysaccharide (K) or lipopolysaccharide (O) antigens are in development. Global *K. pneumoniae* populations display extensive K and O diversity, necessitating a polyvalent vaccine targeted to the serotypes associated with neonatal disease in relevant geographical regions. We investigated the prevalence of K and O types associated with neonatal sepsis in Africa and South Asia to inform maternal vaccine design.

**Methods and Findings:** We analysed 1930 *K. pneumoniae* neonate blood isolates from 13 surveillance studies across 35 sites in 13 countries. We used pathogen whole genome sequencing to predict K and O serotypes and adjust for local transmission clusters, and Bayesian hierarchical meta-analysis to estimate K and O prevalence overall and per region, treating site as a random effect. Eighty-seven K loci were identified. KL2, KL102, KL25, KL15 and KL62 accounted for 49% of isolates. We estimate that 20 K loci, combining the eight most prevalent per region, could cover 72.9% of all infections (95% credible interval: [69.4%, 76.5%]) and ≥70% in each of Eastern, Western and Southern Africa and South Asia. Preliminary findings from three sites suggested sufficient temporal stability of K loci to maintain 20-valent K vaccine coverage over 5-10 years, but more longitudinal data are needed to support this prediction. O types were far less diverse (n=14 types). We estimate the top-5 (O1⍺β,2⍺, O1⍺β,2β, O2⍺, O2β and O4) would cover 86.2% [82.6, 89.9%] of total infections (76–92% per region), while the top-10 would cover ∼99% of infections in all four regions. The main limitations of our study are the reliance on genome sequences to predict K and O serotypes (as serological typing is not available), and a lack of longitudinal data to explore stability of antigen prevalence over time.

**Conclusions:** Neonatal sepsis is associated with diverse K and O types, with substantial geographic and temporal variation even after adjusting for localised transmission clusters. Despite this, a single 20-valent K vaccine could theoretically cover ≥70% of infections in all target regions. Locally-targeted vaccines could achieve higher coverage with lower valency, but are less feasible. In principle, very high coverage could be achieved with lower valency O-based vaccines, however protective efficacy against disease of antibodies targeting the O antigen remains uncertain. Further research is needed on cross-reactivity, antigen exposure and stability of antigens over time, to better inform vaccine development.

## Introduction

An estimated 670,000 newborns die annually from sepsis, with the greatest burden in Africa (∼350,000) and South Asia (∼220,000) [1]. The most common causative agents are the bacterial pathogens *Staphylococcus aureus, Klebsiella pneumoniae* and *Escherichia coli*, with *K. pneumoniae* accounting for >20% of cases [1–4]. The majority of *K. pneumoniae* cultured from neonates are resistant to the antimicrobials recommended by the World Health Organization (WHO) for the treatment of neonatal sepsis [3,5], ampicillin/benzylpenicillin and gentamicin, and it is estimated that nearly 100,000 neonatal sepsis deaths annually are associated with antimicrobial resistant (AMR) *K. pneumoniae* [1]. The Child Health and Mortality Prevention Surveillance (CHAMPS) study, which has sites in seven countries in Africa and Bangladesh in South Asia, identified *K. pneumoniae* as a contributor to 18% of neonatal deaths (and 21% of deaths amongst children under 5 years), with most cultured isolates resistant to ceftriaxone (84%) or gentamicin (80%) [6] the two most commonly used antibiotics.

Given the high burden of neonatal sepsis caused by AMR *K. pneumoniae*, there is an urgent need for preventive measures such as improved infection prevention and control in neonatal healthcare facilities and maternal vaccination [7–9]. It is estimated that a *K. pneumoniae* maternal vaccine given in pregnancy with 70% efficacy administered with coverage equivalent to that of the maternal tetanus vaccine (∼70%) could avert ∼400,000 neonatal sepsis cases and ∼80,000 associated deaths annually worldwide, with the greatest impact in Africa and South Asia [10]. The WHO has identified maternal *K. pneumoniae* vaccine as a ‘critical’ priority, with potential to halve the antibiotic use associated with treatment of *K. pneumoniae* neonatal sepsis globally and save >$US 270 million annually in hospital costs [11], and a WHO Technical Advisory Group has been established to develop a WHO Research Roadmap for vaccines against *K. pneumoniae*.

Polysaccharide antigens, specifically the capsular polysaccharide (K) and lipopolysaccharide (O) antigens, are considered promising candidates for a maternal *K. pneumoniae* vaccine [7]. There are several licensed capsular vaccines against other invasive bacterial pathogens, which can induce protection following a single dose [12–14]. A 6-valent capsular vaccine against group B *Streptococcus* has been developed, and its potential for use in pregnant women to prevent neonatal infection is under investigation [15]. Extensive serotype diversity (>80 K and >10 O) poses a key challenge for *K. pneumoniae* vaccine design. O-based vaccines are attractive due to the smaller number of *K. pneumoniae* O serotypes, are safe and immunogenic [16], but there are currently no such vaccines licensed. Furthermore, some studies suggest O-based vaccines may not be effective at preventing *K. pneumoniae* disease, at least in the presence of certain common K serotypes which have been proposed to exert a masking effect by blocking access of antibodies to the shorter O polysaccharide [16,17]. Multi-valent K-based vaccines, or some combination of K and O, may therefore be required for *K. pneumoniae*. Licensed *Streptococcus pneumoniae* glycoconjugate vaccines target up to 20 capsule types; however, the distribution of vaccine-targeted capsule types varies markedly across geographies, reducing vaccine impact [18–20]. Similar to *S. pneumoniae*, understanding the distribution of *K. pneumoniae* serotypes will be critical to inform the design of vaccines aiming to protect neonates in Africa and Asia.

Serological typing for *K. pneumoniae* is not available outside a few high-income country centres, and the primary tool for profiling K and O serotypes is currently whole-genome sequencing. Serotype predictions can be made based on knowledge of how genetic variation in the K and O antigen biosynthesis loci map to variation in serotype, implemented in the genome-analysis software Kaptive [21]. The genetic determinants underpinning O antigen variation are well understood [22] and include genes located within and outside of the O locus (also detected in Kaptive [23,24]). K antigen biosynthesis is encoded in the K locus, and >160 K loci have been defined on the basis of unique gene content, which maps to unique sugar composition and linkages [22]. Half of these K loci correspond to known serologically distinct reference K serotypes [25]; the others have yet to be fully characterised, but available data support that distinct K loci encode synthesis of K antigens with distinct sugar structures [26–28]. K loci are labelled KL1, KL2, etc, with the numbers corresponding to the antigen structures they encode, which are labelled K1, K2, etc. Comparison of serological data for isolates carrying K loci matching to known reference K serotypes indicates 85% concordance [29].

Available genomic surveillance data suggest that *K. pneumoniae* serotypes associated with bloodstream infection differ geographically and temporally [2,30–32]. The limited data available specifically for neonates suggest marked differences in serotype prevalence between sites [2,30], skewed by localised clustering that likely reflects local outbreaks within the neonatal units as occurs frequently in Africa and Asia [4,30,33].

Here, we analyse *K. pneumoniae* isolated from the blood of neonates with suspected sepsis in African and South Asian countries sourced from 13 different prospective surveillance studies over the last decade, aiming to: (1) estimate the prevalence of genomically-predicted K and O serotypes amongst *K. pneumoniae* associated with neonatal sepsis in low- and middle-income countries (LMIC) in Asia and Africa during this time period; and (2) estimate the cumulative coverage of neonatal sepsis cases offered by different sets of K or O antigens, including subgroup analyses assessing coverage within (i) geographical regions, (ii) neonates who died during follow-up, and (iii) infections caused by extended-spectrum beta-lactamase (ESBL) producing or carbapenemase producing (CP) strains. We use whole-genome sequencing to predict K and O serotypes and correct for local transmission clusters, and Bayesian hierarchical meta-analysis to estimate total and regional serotype prevalence and theoretical vaccine coverage. Our findings will inform the design and deployment of maternal vaccines against *K. pneumoniae* neonatal sepsis.

## Methods

### Ethical considerations

Each contributing study obtained local ethical approval, listed in **Table S1**. The Baby GERMS-SA [34] and MLW [30] studies were granted consent waivers from local ethics committees for the use of routine diagnostic specimens/isolates and clinical data for the research; all other studies obtained written informed consent from the parents or guardians of participating neonates. Approval for the meta-analysis presented here was granted by the Observational / Interventions Research Ethics Committee of the London School of Hygiene and Tropical Medicine (ref #29931). Anonymised data from each primary study were shared for analysis, including date of specimen collection, hospital site identifier (where the study included more than one site), and mortality outcome (where available) for each participant, along with pathogen genome data for the corresponding bacterial isolate.

### Bacterial isolates and sequence analysis

Whole-genome sequences of *K. pneumoniae* isolated from neonates in low- and middle-income countries in Asia and Africa were sourced from prospective clinical studies of neonatal sepsis (BARNARDS [2], SPINZ [35], Baby GERMS-SA [34], MBIRA [36], DH [37], GBS-COP [38], NIMBI-plus, NeoOBS-India [3], NeoBAC), long-term prospective surveillance of bloodstream infection (MLW [30], KWTRP, CHRF [39]), and referral-based surveillance of blood cultures in Pakistan (AKU) (**Table S2**).

For the prospective studies (BARNARDS, SPINZ, Baby GERMS-SA, MBIRA, DH, GBS-COP, NIMBI-plus, NeoOBS-India, NeoBAC), blood culture was performed for all neonates with clinically suspected sepsis, and all blood culture isolates identified as *Klebsiella* that could be later revived for DNA extraction were included in sequencing and this meta-analysis. Baby GERMS-SA, GBS-COP, NeoOBS, KWTRP and MLW also included *Klebsiella* cultured from cerebrospinal fluid (CSF) of participants.

For the long-term prospective surveillance, isolates from all positive blood cultures (CHRF, KWTRP, MLW) or CSF cultures (KWTRP, MLW) were stored for future research, and all those identified as *Klebsiella* isolated from neonates that could be later revived for DNA extraction were included in sequencing and this meta-analysis. At KWTRP, all neonates had blood culture on admission and again if their clinical condition deteriorated. At MLW, cultures were performed for all neonates with clinical signs of sepsis or meningitis, temperature >37.5 °C, or other signs of clinical deterioration. At CHRF, blood cultures were performed as part of routine diagnostics whenever clinically indicated.

For the AKU referral-based study, all positive blood cultures obtained from infant inpatients (including emergency) at Aga Khan University Hospital (AKUH), as well as all those obtained from the AKUH Laboratory Network (comprising laboratories across Pakistan), as part of routine diagnostics were stored for analysis. All those identified as *Klebsiella* isolated from neonates during the study period, that could be later revived for DNA extraction, were included in sequencing and this meta-analysis.

Details of microbiology, sequencing and bioinformatics methods used in each study, including quality-control criteria applied prior to sharing genome sequences for this meta-analysis, are summarised in **Table S2**. All studies utilised Illumina platforms and assembled genomes using SKESA v2.3.0 (MBIRA) or SPAdes (all other studies). Genome assemblies were analysed using Kleborate v3.0 [40] to confirm species, identify multi-locus sequence types (STs), antimicrobial resistance (AMR) and hypervirulence determinants, and to identify K and O types via Kaptive v3.0 [21]. For the K antigen, we used the K locus call as the unit of analysis. For the O antigen we used O type as the unit of analysis, using the latest O subtype nomenclature [22], inferred from the combination of O locus and O type reported by Kaptive v3.0 (full genetic definitions in **Appendix S1**). Genomes identified by Kaptive as untypeable (<1%) were included in the denominators for all analyses.

### Inclusion criteria for meta-analysis

Inclusion criteria for individual samples from these studies were: *K. pneumoniae* isolated from the blood or CSF of a neonate (defined as 0-30 days post birth, or 0-28 days for Baby GERMS-SA, DH, MBIRA, MLW, and SPINZ), between 2013-2023. CSF isolates were included from four studies only (NeoOBS-India, n=9; KWTRP surveillance, n=5; GBS-COP, n=2; MLW, n=31). Repeat isolates of *K. pneumoniae* from the same participant with the same KL were excluded, such that each genome represents a unique infection from an individual neonate. Genome sequences identified as species other than *K. pneumoniae*, or failing to meet the meta-analysis quality control criteria for assemblies (≤1000 contigs and genome size 5-6.2 Mbp, as reported by Kleborate v3), were excluded. Individual study sites with fewer than 10 high-quality *K. pneumoniae* genomes passing the inclusion criteria were excluded (81 genomes from 21 sites). A flow diagram for sample inclusion is shown in **Figure S1**, including available information on the number of samples that were not stored, were unable to be revived for DNA extraction, failed sequencing, or failed genome quality control for each contributing study.

### Subgroup definitions

African countries were assigned to regions using the United Nations Statistics Division standard M49 [41] (intermediate region level). Bangladesh, India and Pakistan were assigned to South Asia.

Isolates were assigned as ESBL if they had a Kleborate v3 resistance score of ≥1 (indicating detection of at least one known ESBL gene in the genome), and as CP if they had a Kleborate v3 resistance score of ≥2 (indicating detection of at least one known carbapenemase gene in the genome [40]).

Fatalities were defined as all neonates with blood-culture isolates of *K. pneumoniae* who were recorded as dying from any cause, either during the hospital admission in which the blood culture was taken or during a defined follow-up period, depending on the study. Some studies recorded in-hospital deaths only (Baby GERMS-SA, KWTRP, NeoBAC, SPINZ), others also undertook follow-up for a defined period (MBIRA: 30 days post-enrolment, NeoOBS-India: 28 days post-enrolment, DH: 28 days of life, BARNARDS: 60 days of life). Mortality data were not available from the MLW, CHRF, NIMBI-plus and AKU surveillance studies.

### Identifying clusters

To ensure a simple and reproducible analysis, we used pairwise single nucleotide variant (SNV) distances downloaded from Pathogenwatch (which we refer to as ‘PW distances’). These are calculated across the core gene library defined by Pathogenwatch for *K. pneumoniae* (1,972 genes, 2 ,172, 367 bp) [42]. There is emerging consensus across multiple studies that a threshold of 21-25 genome-wide SNVs (estimated using the more common method of mapping reads to a complete *K. pneumoniae* reference genome) is suitable for identifying nosocomial transmission clusters of *K. pneumoniae* [43–45]. We recently showed that a genome-wide threshold of 25 SNVs is equivalent to a PW distance of 10 SNVs in *K. pneumoniae* [46], hence we used a threshold of PW distance ≤10 SNVs, between genomes isolated from the same study site and within 4 weeks (28 days), to define putative nosocomial transmission clusters for our primary analysis. We explored sensitivity of our analyses to clustering thresholds by varying the genetic distance (1, 2, 3, 5, 10, 15, 20, 25 or 50 PW SNVs) and temporal distance (0, 1, 2, 4, 8, 52 weeks) (reported in **Appendix S2**). Clustering was done in R using the *igraph* package (v2.0.3) [47], by first creating an (undirected) graph from a list of edges connecting isolates from the same site and meeting the genetic and temporal distance thresholds, then extracting clusters (groups of linked components, i.e. single-linkage clustering).

### Statistical analysis

Crude prevalence for a given K locus (or O type) was calculated as the proportion of genome counts (crude prevalence), or proportion of unique clusters (cluster-adjusted crude prevalence), carrying that locus/type.

Global (i.e. across all included samples) and regional prevalences were estimated using a Bayesian framework to fit a generalized hierarchical linear mixed model with a binomial family, with events being the count (raw or cluster-adjusted) of isolates with each locus observed at each site, with fixed effects for locus and region and random effects for site and locus|region. Specifically, the data comprises *N* sites and *K* loci resulting in *K*N* observations. One model each was fitted across all locus types and sites, for each combination of raw and cluster-adjusted K and O counts (4 models in total). Models were fitted using the *brms* package in R (v2.22.0) [48], which provides an interface to the probabilistic programming language Stan (v2.32.2) [49].

Model specification is shown below. Each grouping *i* represents a unique combination of locus, site and region.

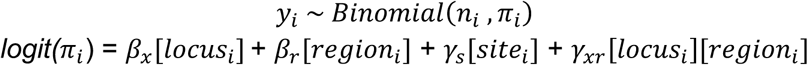

The priors for the fixed effects (*site* and *region*) were a Uniform distribution from -∞ to ∞ (default *brms* priors). For the standard deviations of the random effects (for *site* and *locus:region*), priors were specified as student-t distributions with 1 degree of freedom, location 0, and scale 10. Each model was fit with 4 chains, each running for 6000 iterations, with 3000 burn-in iterations. Control parameters were set to ensure stable sampling, with adapt_delta = 0.999999 and max_treedepth = 55. Model diagnostics (Rhat, effective sample size ratio, and divergent transitions) were inspected for all runs (see **Appendix S3**). In cases where initial model diagnostics warranted, models were run again with 30,000 iterations with 15,000 burn-in iterations.

To estimate global and regional prevalence for subgroups (fatalities, ESBL, or CP), the primary data were filtered to include only cases belonging to that subgroup, then any sites with fewer than 10 included cases were excluded from the analysis, before fitting the same model specified above.

For each model we used the *posterior_epred* function from the *brms* package to generate a posterior distribution of predicted values from the fitted model, for each *locus* (i.e. the *locus* effect, reflecting global estimate) and for each combination of *locus* and *region* (i.e. the *locus + region + locus:region* effects, reflecting the regional estimates for each locus). These were summarised to obtain the mean value and 2.5% and 97.5% quantiles, which we interpret as the point estimate and 95% credible interval (CI) for the global and regional prevalences of each locus/type.

To estimate cumulative coverage for theoretical vaccine analyses, for each set of K loci (or O types) we obtained posterior distributions by summing the relevant locus/type estimates for each draw, then calculated the mean value and 2.5% and 97.5% quantiles, which we interpret as the point estimate and 95% CI for cumulative sets of loci/types.

Leave-one-out analysis was undertaken to explore robustness of global cluster-adjusted prevalence estimates, and particularly to understand how global prevalence estimates are influenced by individual studies. The above Bayesian models were each run repeatedly using the same methods outlined above (using cluster-adjusted counts), but excluding one study each time, and loci/types were ranked according to the resulting prevalence estimate.

To assess whether the prevalence of common K loci were associated with year or region of sampling, for each K locus in the overall top-30 we used the *logistf* R package (v1.26.0) to fit a logistic regression using Firth’s bias reduction method, with the response variable being 1 or 0 (to indicate the presence of this locus vs any other) and predictors being year and region. A p-value <0.05 was used to assess significance.

All data processing, analyses and data visualisations were conducted using R Statistical Software (v4.4.1) [50] with packages *dplyr* (v1.1.4), *tidyverse* (v2.0.0), *ggplot2* (v3.5.1), *ggridges* (v0.5.6), and *patchwork* (v1.3.0).

### Code availability

Model diagnostics are shown in **Appendix 3**. Model input, output and code, and all other analysis and plotting code required to produce all figures and tables, including supporting items and appendices, are available at <u>github.com/klebgenomics/KlebNNSsero</u> (DOI: 10.5281/zenodo.15756796).

### Data availability

Sample-level data, including K/O calls, accessions and source information are given in **Table S3**. Raw and adjusted counts data used for modelling, and model code and outputs, are available at <u>github.com/klebgenomics/KlebNNSsero</u> (DOI: 10.5281/zenodo.15756796). An interactive web application, implemented in R shiny, is also available to (i) reproduce and explore the modelled estimates for prevalence and coverage data included in this paper, and (ii) undertake additional analyses of the raw data (e.g. to explore crude pooled estimates of prevalence and coverage for different subsets of loci and/or different subsets of samples). The web application is available at https://klebsiella.shinyapps.io/neonatal/. Raw whole-genome sequence data were deposited by the originating study teams in INSDC databases, under the following BioProjects: BARNARDS, PRJEB33565; SPINZ, PRJEB46513; MLW, PRJEB42462; NIMBIplus, PRJNA1168993; DH, PRJEB70311; BabyGERMS, PRJNA796486 and PRJNA1282934; CHRF, PRJEB90555; NeoOBS-India, PRJEB70311; GBS-COP, PRJNA1175467; Kilifi, PRJNA1265413; MBIRA: PRJNA1274034, NeoBAC, PRJNA1265413; AKU, PRJNA126164.

## Results

A total of 1930 *K. pneumoniae* genomes, each representing a unique neonatal infection sampled in the last decade, were included from 35 sites in 13 countries from four regions in Africa and Asia (**Table 1**, **Figures 1 and S1**). Four studies included CSF isolates, accounting for n=47 infections (2.4%). Nearly all genomes yielded typeable K and O biosynthesis loci (99.1% K, 99.5% O), and a total of 87 distinct K loci and fourteen O types were identified across the studies.

**Figure 1.**
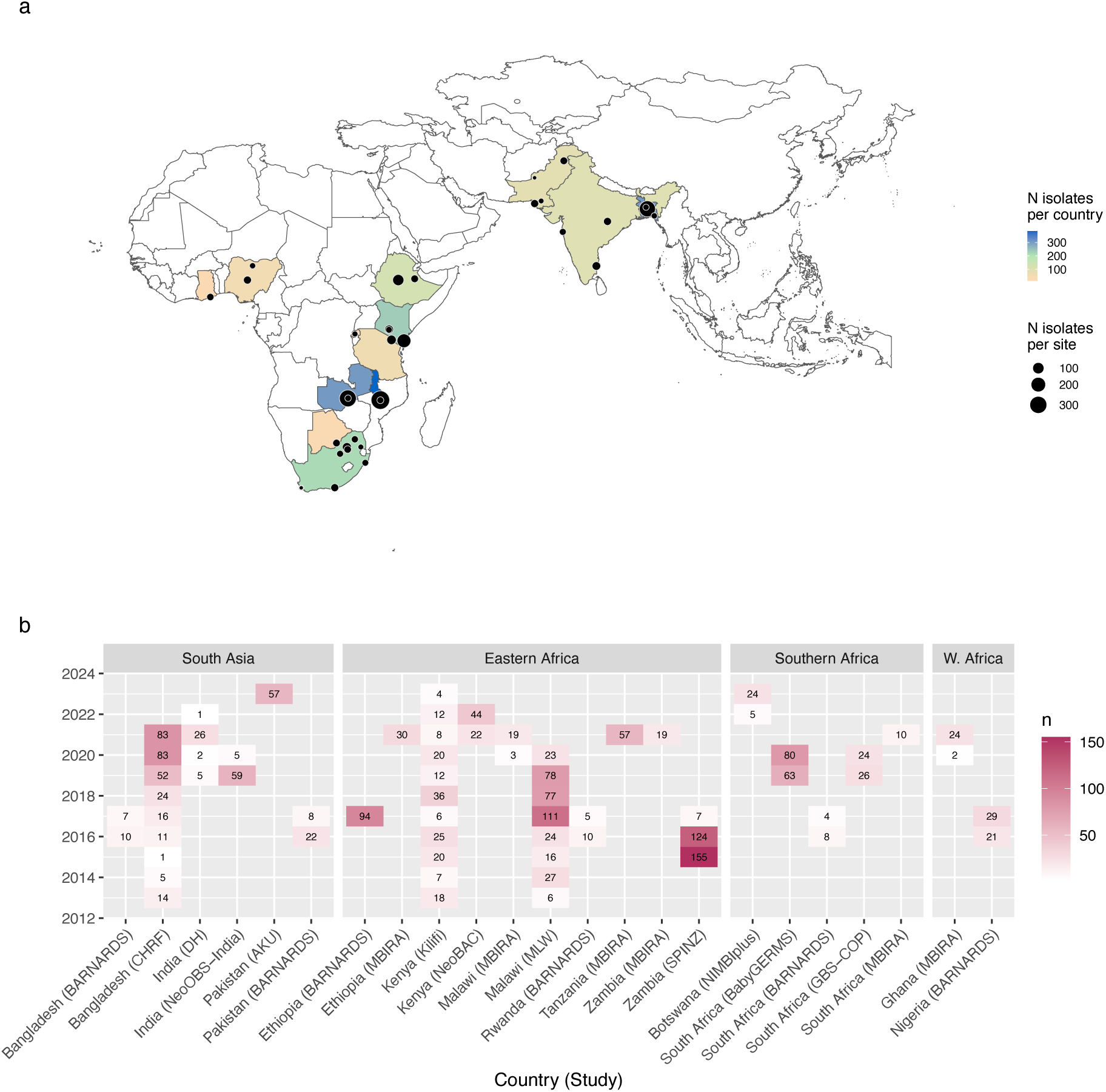
Geotemporal distribution of isolates included in the meta-analysis. (a) Barplots indicate the number of samples included per year, from each study site (labelled as “Country, Study”), stratified by region. (b) Map shows location of study sites. Each point represents a study site, sized to indicate the number of samples included from that site, as per inset legend. Countries are coloured to indicate the total number of isolates included from that country, as per inset legend. Base map source: Natural Earth, https://www.naturalearthdata.com/downloads/10m-cultural-vectors/, accessed using rnaturalearth R package v1.10, terms of use: https://www.naturalearthdata.com/about/terms-of-use/.

**Table 1.** Details of studies and sites included in the meta-analysis.

| Region | Country | Study | Unique Sites | Unique Isolates | Period | Unique K loci | Unique O types |
| --- | --- | --- | --- | --- | --- | --- | --- |
| Eastern Africa | All | - | 11 | 1119 | - | 60 | 11 |
|  | Ethiopia | BARNARDS | 1 | 94 | 2017-2017 | 11 | 4 |
|  |  | MBIRA | 1 | 30 | 2021-2021 | 10 | 6 |
|  | Kenya | Kilifi | 1 | 168 | 2013-2023 | 43 | 10 |
|  |  | NeoBAC | 2 | 66 | 2021-2022 | 5 | 3 |
|  | Malawi | MBIRA | 1 | 22 | 2020-2021 | 11 | 4 |
|  |  | MLW | 1 | 362 | 2013-2020 | 35 | 10 |
|  | Rwanda | BARNARDS | 1 | 15 | 2016-2017 | 6 | 3 |
|  | Tanzania | MBIRA | 1 | 57 | 2021-2021 | 5 | 3 |
|  | Zambia | MBIRA | 1 | 19 | 2021-2021 | 10 | 6 |
|  |  | SPINZ | 1 | 286 | 2015-2017 | 13 | 7 |
| Southern Africa | All | - | 10 | 244 | - | 33 | 8 |
|  | Botswana | NIMBIplus | 1 | 29 | 2022-2023 | 8 | 7 |
|  | South Africa | BARNARDS | 1 | 12 | 2016-2017 | 8 | 4 |
|  |  | Baby GERMS-SA | 6 | 143 | 2019-2020 | 24 | 8 |
|  |  | GBS-COP | 1 | 50 | 2019-2020 | 12 | 6 |
|  |  | MBIRA | 1 | 10 | 2021-2021 | 6 | 3 |
| Western Africa | All | - | 3 | 76 | - | 27 | 9 |
|  | Ghana | MBIRA | 1 | 26 | 2020-2021 | 14 | 7 |
|  | Nigeria | BARNARDS | 2 | 50 | 2016-2017 | 18 | 8 |
| South Asia | All | - | 11 | 491 | - | 63 | 12 |
|  | Bangladesh | BARNARDS | 1 | 17 | 2016-2017 | 12 | 8 |
|  |  | CHRF | 3 | 289 | 2013-2021 | 41 | 10 |
|  | India | DH | 1 | 34 | 2019-2022 | 8 | 8 |
|  |  | NeoOBS | 2 | 64 | 2019-2020 | 26 | 9 |
|  | Pakistan | AKU | 3 | 57 | 2023-2023 | 26 | 9 |
|  |  | BARNARDS | 1 | 30 | 2016-2017 | 9 | 6 |
| Total | - | - | 35 | 1930 | - | 87 | 14 |

The raw distribution of K loci is shown in **Appendix S1.1**. Four K loci were common, with crude prevalence exceeding 5% each (KL102, KL15, KL2, KL25), and together accounting for 45% of isolates. The distribution of K loci varied across countries, with no K loci found in all countries (KL2 and KL25 seen in n=11 of 13 countries), and only twelve K loci found in all four regions sampled (KL2, KL3, KL10, KL15, KL17, KL19, KL25, KL39, KL48, KL62, KL102, KL122; accounting for 59% of isolates). Most K loci were observed in combination with one or two O types, and these patterns were associated with the clonal population structure of *K. pneumoniae* and spatiotemporal clustering (represented by lineage, i.e. sequence type (ST) in **Appendix S1.1**). In some countries the samples were dominated by specific KL/O combinations; these were mostly associated with a single lineage or ST, and identified in temporal clusters at a single study site (**Appendix S1.2**), consistent with local transmission in the hospital or the wider community. For example, isolates from Ethiopia were dominated by ST35-KL108/O1⍺β,2β (30%) and ST37-KL15/O4 (22%), but these were all isolated from the same hospital in 2017 and these clones were not detected in a later study at a different site in Ethiopia. Similarly, isolates from Zambia were dominated by ST307-KL102/O2β, all isolated from one hospital in 2015-2017, but this clone was not identified in a later study from a different hospital in Zambia. In Tanzania, all data came from a single study site in 2021, and 90% were ST1741-KL104/O1⍺β,2β; however this clone was rare elsewhere (n=5, 5.6% in India; n=5, 2.1% in Kenya; not detected elsewhere). The genome data therefore supports that observed K (and O) locus prevalence can be heavily influenced by localised nosocomial outbreaks, as noted above.

### Modelled K locus prevalence estimates

We used Bayesian modelling to estimate global and regional prevalence for each K locus, based on meta-analysis of the 1930 *K. pneumoniae* genomes included from 13 studies in four regions (**Figure 2**). Given the influence of localised outbreak events on observed K locus counts at each site, we used cluster-adjusted counts whereby each genetic-temporal cluster (≤10 SNVs and ≤28 days) within a site (i.e. the same neonatal unit) is treated as a single observation (see **Methods**). Five loci (KL2, KL102, KL25, KL15, KL62) had mean prevalence estimates exceeding 4% each (with lower 95% credible interval (CI) each ≥3.2%, see **Figure 2a**). A further seven K loci (KL30, KL10, KL17, KL23, KL51, KL112, KL39) had mean estimates exceeding 2% each (lower CI, ≥1.3%). In total, 27 K loci had mean prevalence estimates exceeding 1% (see **Figure 2a**).

**Figure 2.**
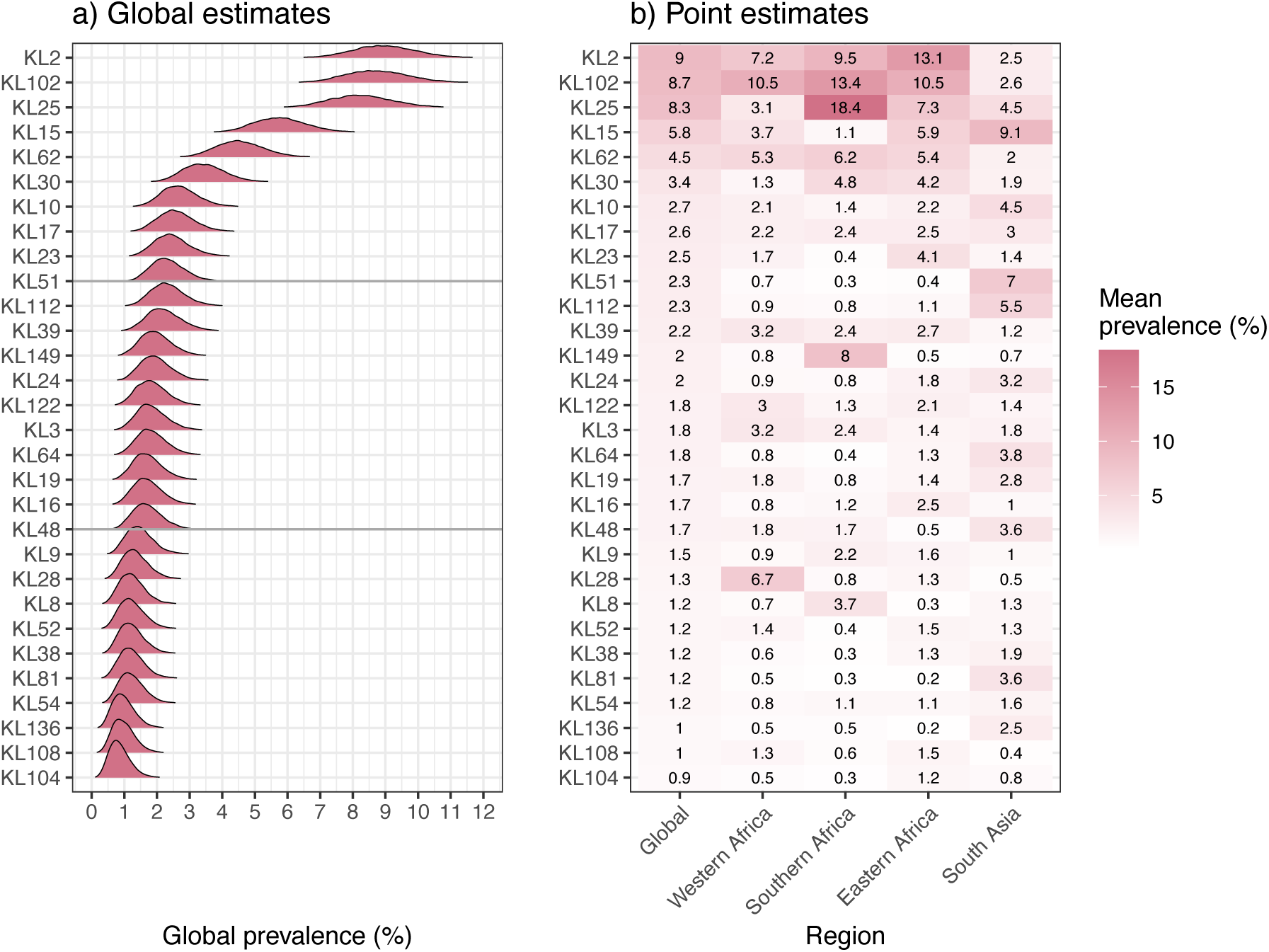
Modelled estimates of cluster-adjusted global and regional prevalence of K loci. (a) Posterior density distribution for global prevalence estimates (i.e. across all sites), modelled using cluster-adjusted counts per site, for top-30 K loci ordered by the mean point estimate. Horizontal lines indicate the top 10 and 20 loci. (b) Regional prevalence estimates. Cells are coloured and labelled to indicate the mean cluster-adjusted regional prevalence estimates for each K locus in each region, according to the inset legend. The underlying posterior distributions for K locus prevalence per region are shown in **Figure S5**.

K locus prevalence estimates varied substantially across regions (**Figures 2b, S2, S3**). Some of the global top-30 loci had significantly higher prevalence in a single region (**Figure S3**), e.g. KL149 in Southern Africa (8.0% [95% CI 4.3%,12.7%], vs ≤0.84% in other regions), and KL15 and KL51 in South Asia (9.1% [5.9%, 13.0%] vs ≤6% elsewhere and 7.0% [4.2%, 10.6%] vs ≤0.7%, respectively) (**Figures 2b, S3**). Outside the top-20, KL81 was common in South Asia (3.6%, [1.6–6.3%]) but very rare elsewhere (≤0.5%), and KL28, KL116 and KL53 were common in Western Africa (6.7% [2.0%, 14.0%], 5.6% [1.4%, 12.9%] and 4.0% [0.7%, 10.0%], respectively) but were rarely detected elsewhere (0.3–1.4%; see **Figures S2**).

These regional differences could not be explained by differences in the timing of samples from different regions (**Appendix S1.3**). In a simple logistic regression for each K locus in the top-30, with year and region as linear predictors, region was significantly associated with 15 K loci (^ in **Figure S3**) but year was not significant for any K loci.

### Theoretical K antigen coverage

To estimate the proportion of neonatal sepsis cases that could theoretically have been covered by a vaccine comprising subsets of K antigens, we modelled cumulative coverage based on prevalence estimated from raw (observed) counts (**Figure 3**). We estimate a vaccine with the global top-20 K-loci would cover 72.8% of all cases [95% CI 69.3%, 76.4%], with higher coverage in Eastern Africa (75.7% [71.1%, 80.6%]) and Southern Africa (76.4% [66.6%, 86.8%]) and lower coverage in Western Africa (50.5% [36.9%, 66.4%]) and South Asia (67.8% [61.1%, 75.0%]) (**Table S4**). The marginal gain in coverage associated with adding more loci (in order of global rank) reduced substantially beyond the first 5 K-loci; these would provide theoretical global coverage of 48.8% of cases [46.0%, 51.7%], but we estimated the next five loci (global ranks 6-10) would provide additional global coverage of just 9.8% [9.6%, 10.2%], a further five (global ranks 11-15) 8.2% [8.0%, 8.4%], and a further five (global ranks 16-20) 5.8% [5.7%, 6.0%] (**Table S4**, **Figure 3**).

**Figure 3.**
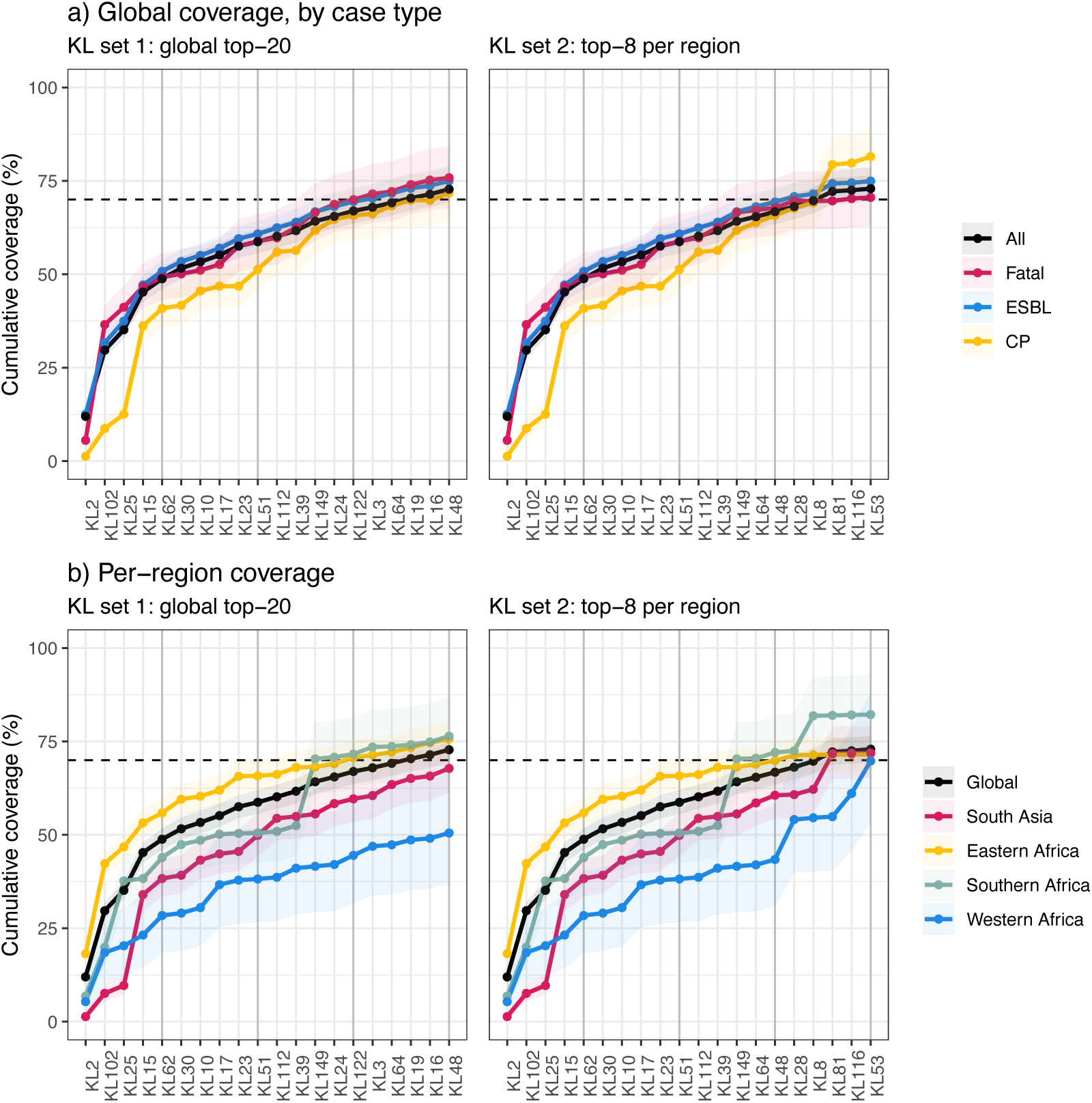
Cumulative coverage estimates for sets of K loci. (a) Global (overall) coverage of neonatal sepsis and subgroups of sepsis cases (fatal, ESBL, CP); and (b) per-region coverage of neonatal sepsis; selecting either the top-20 K loci ordered by cluster-adjusted global prevalence estimate (KL set 1) or selecting the top-8 K loci within each region (top-7 in Southern Africa) (KL set 2, total 20 K loci). KL, K locus; ESBL, extended-spectrum beta-lactamase; CP, carbapenemase-producing.

The global top-20 K loci provided very similar estimated coverage of neonatal fatalities (outcome data available for n=1063, 55.0% of genomes; of which n=394, 37.1% died), and coverage of infections with ESBL-producing strains (n=1747, 90.5% of genomes overall, 78.3–96.7% per region) (see **Table S4**, **Figure 3**). Infections with CP strains were very common in South Asia (87% in Bangladesh, 78% in India, 87% in Pakistan); common in Nigeria (46%), Ghana (27%) and South Africa (31%); and rare elsewhere (<3%). Estimated coverage of these CP infections was similar to the overall coverage (top-20 K loci covering 71.7% [64.8–79.0%]). However, four of the global top-5 K loci (KL2, KL102, KL25, KL62) were rare amongst CP isolates, and so the cumulative coverage of the top-5 was low for CP infections (40.9%, compared with 48.8% of total infections and 50.8% of ESBL infections).

As coverage estimates using the global top-20 K loci varied substantially by region (**Table S4, Figure 3**), we explored alternative strategies to select K loci in order to achieve better regional coverage. For each region, a locally tailored set of K loci including the top 20 most prevalent in that region could provide ≥80% [lower CI, ≥62%] coverage in the target region (**Figure S4**). However, any such formulations would provide insufficient coverage of non-target regions (**Figure S4**). We estimate an alternative set of 20 K-loci, comprising all those that ranked in the top-8 for any region, would cover ≥69.8% of cases in each region (range 69.8-82.2% per region, 72.9% overall), with similar coverage of ESBL, CP and fatalities (74.9%, 81.5%, 70.6%, respectively; see **Figure 3**, **Table 2**).

**Table 2.**
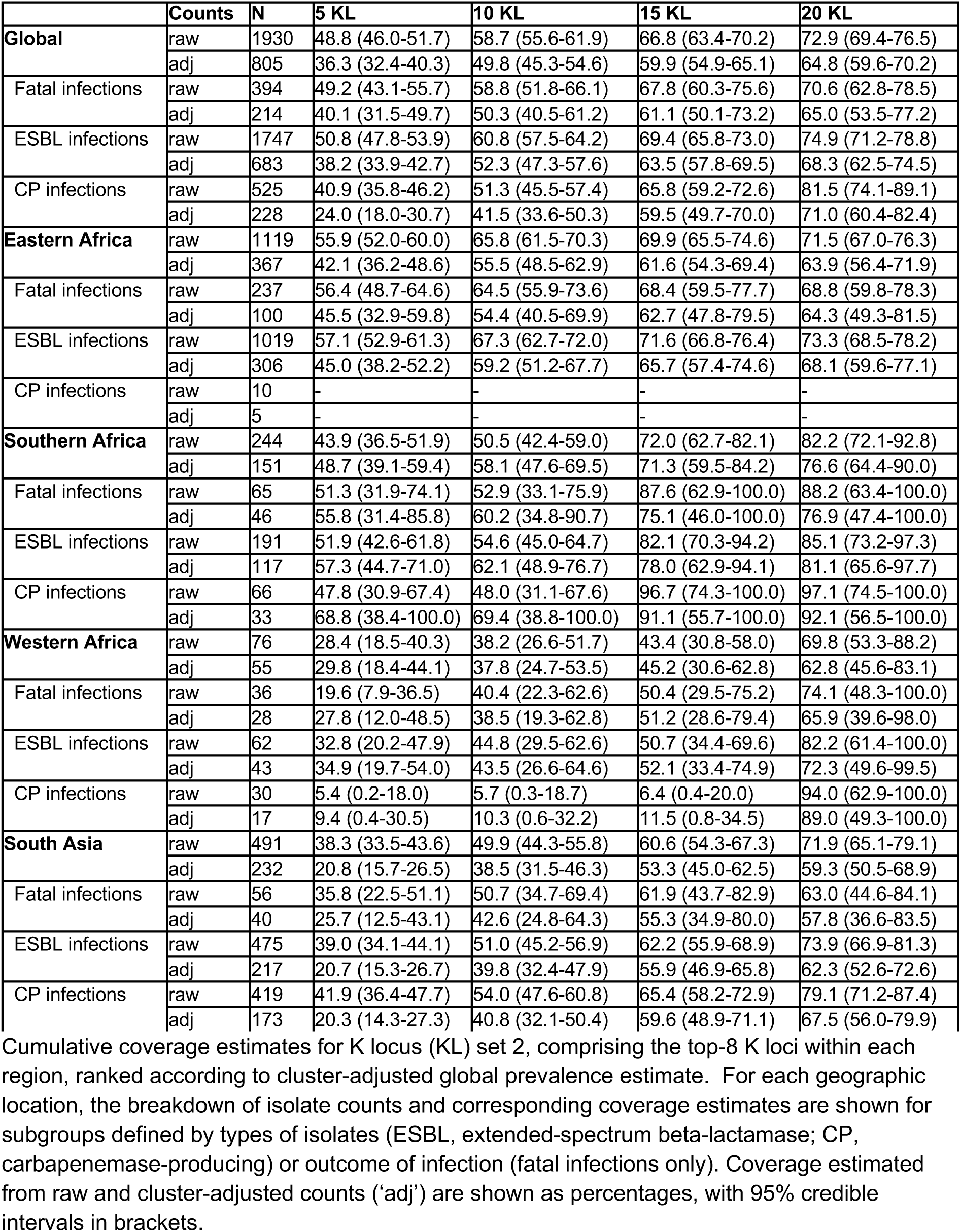
Coverage estimates for 20 loci including the top-8 per region.

### O type prevalence and coverage

Global prevalence modelling of predicted O types (**Figure 4a**) confirmed the population was dominated by two O1 subtypes, O1⍺β,2⍺ and O1⍺β,2β. The median prevalence estimates for these were 26.7% [95% CI 23.6%, 29.9%]) for O1⍺β,2⍺, and 21.8% [19.1%, 24.8%]) for O1⍺β,2β. Next most common (≥9% each) were O2β (15.8% [13.5%, 18.4%]) and O4 (9.1% [7.2%, 11.1%). Combined, these top-4 O types were estimated to cover 81.1% [77.6%, 84.6%] of all included cases. Five other O types were detected with >3.0% prevalence, including O2⍺ (6.2% [4.6%, 7.9%]); two O3 subtypes (O3γ, 6.0% [4.5%, 7.8%] and O3⍺/O3β, 2.8% [1.8%, 4.1%]), O5 (5.4% [3.9%, 7.0%]) and O13 (4.9% [3.5%, 6.5%]). Loci associated with other O types were very rare (mean estimates <0.4%, upper 95% Cis <0.9%; see **Figure 4a**).

**Figure 4.**
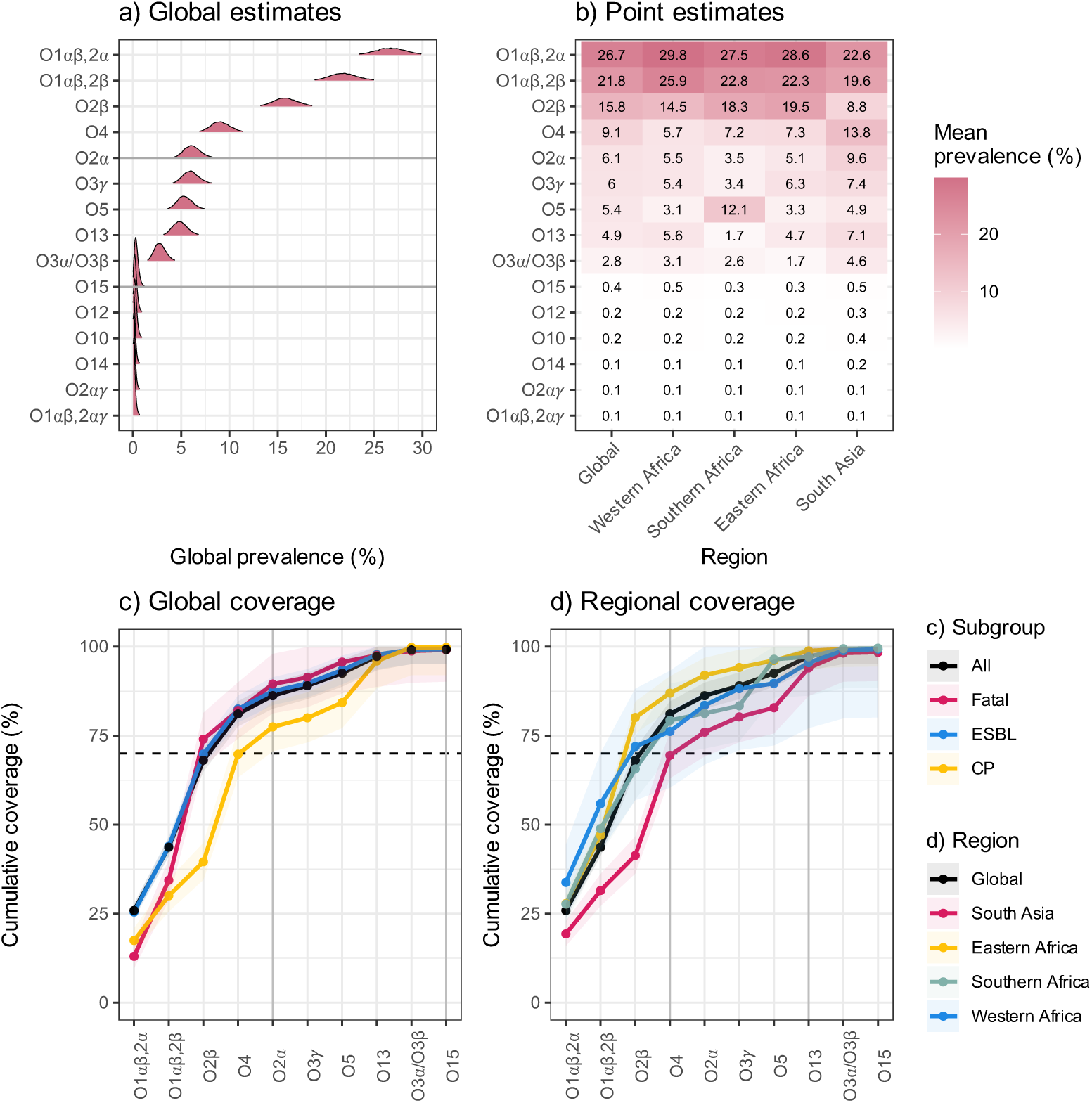
Modelled estimates of cluster-adjusted global and regional prevalence and coverage of O types. (a) Posterior density distribution for global (overall) prevalence estimates, modelled using cluster-adjusted counts per site, for O types ordered by the mean point estimate. Horizontal lines indicate the top 5 and 10 types. (b) Regional mean prevalence estimates. Cells are coloured and labelled to indicate the cluster-adjusted regional prevalence estimates for each O type in each region, according to the inset legend. (c) Global (overall) coverage of neonatal sepsis and subgroups of sepsis cases (fatalities, ESBL; extended-spectrum beta-lactamase, CP; carbapenemase-producing), and (d) per-region coverage of neonatal sepsis, selecting the top-10 O types ordered by cluster-adjusted global prevalence.

There was some regional variation in O types (**Figures 4b and S5, Table S5**). Whilst O1 variants dominated in all regions, O2β was consistently the third most common in African regions (14.5–19.5%) but was less common in South Asia (8.8%), which had higher prevalence of O2⍺ (9.6%, vs 3.5–5.5% in African regions) and O4 (13.8%, vs 5.7–7.3% in African regions). Compared with other regions, Southern Africa had higher prevalence of O5 (12.1%, vs 3.1–5.0% elsewhere), and lower prevalence of O3γ (3.3%, vs 5.4–7.4% elsewhere) and O13 (1.7%, vs 4.7–7.1% elsewhere). As with K locus prevalence, there was no evidence that regional differences were driven by differences in time of sampling (i.e. the calendar year of sampling for different sites); e.g. in South Asia, the prevalence of O2β was persistently lower than elsewhere, and O4 and O2⍺ persistently higher, across the combined sampling period (**Appendix S1.4**). The differences in O types were also not explained by localised clonal outbreaks; e.g. in South Asia O4 was associated with 15 different ST-KL combinations (the most common being ST11-KL15 found in three sites in Bangladesh and two in India) and O2⍺ with nine ST-KL combinations (**Appendix S1.5**). O5 in Southern Africa was primarily associated with a single clone ST17-KL25/O5, but this was distributed across seven sites in South Africa and one in Botswana, consistent with endemic spread in the region.

The estimated coverage of infections per subgroup and region are shown in **Figure 4c-d, Table S5**. We estimate the top-5 O types (O1⍺β,2⍺, O1⍺β,2β, O2β, O2⍺, O4) collectively cover 86.2% [95% CI 82.6%, 89.9%] of total cases, 89.5% [81.3%, 98.0%] of fatalities, 87.5% [83.6%, 91.4%] of ESBL and 77.5% of CP [70.6%, 84.5%]; and ≥76% of cases in each region (range 76.0-92.0% per region, lower CI ≥69%) (**Table S5**).

### Sensitivity analysis

To explore the sensitivity of our analyses to which primary studies were included, we repeated the meta-analysis leaving out each of the n=13 studies in turn (**Appendix S2.1-2.2**). Global modelled ranks for K loci were robust to study inclusion, with the same top-5 in all analyses and the top-20 also quite stable (only 5 loci ranking top-20 in any leave-one-out analysis were not included in the overall top-20). The cumulative coverage estimated for the final global top-20 K loci was also very robust to study inclusion (with the exception of regional coverage in West Africa, for which we had just two contributing studies, see **Appendix S2.1**). O type rankings were also very robust to study inclusion, with the same top-4 for all analyses, and cumulative coverage estimates for the top-4 were stable across analyses (see **Appendix S2.2**).

The impact of cluster-adjustment on the modelled and simple weighted global prevalence estimates for K loci are shown in **Appendix S2.3–2.4**. Note that cluster-adjustment is equivalent to counting only the index case for each cluster in each site, removing the random effect of nosocomial transmission events that may be driven by local infection control issues rather than specific bacteriological properties. Cluster-adjustment had a substantial effect (0.5–9.0% reduction in mean global prevalence estimate) for eight K loci, due to raw counts of these K loci each being inflated by a few large clusters (1-3 clusters per K locus accounting for 54-100% of total observations for that K locus). The largest (≥3.0% prevalence) reductions were observed for the three loci with highest crude count (KL2, KL102 and KL15), although these remained in the top-4 even after cluster-adjustment (see **Appendix S2.4**). The next most affected by cluster-adjustment were KL104, KL157, KL81, KL108, and KL149 (0.5–2.5% reduction); these were ranked 6, 16, 9, 11, 8 based on raw counts but only KL149 was in the top-20 following cluster-adjustment (the other four being ranked 30, 73, 26 and 29, respectively). We also explored sensitivity to the choice of genetic and temporal thresholds used for clustering, and found only minor impact on per-locus prevalence estimates and ranks (**Appendix S2.5–2.7**).

The primary estimates of cumulative coverage reported above were based on raw counts (i.e. observed infections). This is because we do not expect that a vaccine would prevent nosocomial transmission clusters in neonatal units, as these most often represent point source outbreaks due to contaminated equipment or hospital environment, rather than neonate-to-neonate in a transmission chain where prevention of an index case would prevent onward transmission to others. For comparison, estimates of K or O coverage based on cluster-adjusted prevalence are shown in **Appendix S2.8-2.10 and Tables 2, S4, S5**. Overall the cluster-adjusted coverage estimates were a few percentage points lower, as clusters were often associated with common serotypes, although this varied by region.

Our data set is not suitable to assess sensitivity to time of sampling (i.e. the calendar year of sampling for different studies), or to investigate changes in locus prevalence over time within sites, since the contributing studies were conducted in different time periods (see **Figure 1**) and year of isolation is confounded with both study and geographical location. However, to motivate future longitudinal studies addressing this issue, we explored the data from three sites (in Bangladesh, Malawi, and Kilifi) where blood culture isolates had been routinely collected over extended periods. Ranking K loci and O types based on simple pooled prevalence estimates across these three sites from isolates collected up to 2017 (n=512 isolates), the top-20 K loci account for 69.9% (66.9–73.6% per site) of neonatal sepsis isolates collected after 2017 (n=512), and the top-5 O types for 80.3% (73.98–90.4% per site) collected after 2017 (**Figure S6**).

## Discussion

In this study we estimated the prevalence of K loci and O types among *K. pneumoniae* causing neonatal sepsis in Africa and South Asia, overall and for each of the four included regions (**Figures 2, 4**). KL2, KL102 and KL25 were the most common K loci, with cluster-adjusted prevalence of 8.3–8.9% each, and together covering 35% of neonatal sepsis isolates (**Figures 2, 3**). Two other K loci, KL15 and KL62, each exceeded 4.5% prevalence, and together with the top three loci accounted for 49% of isolates (**Table 2**).

Our analyses revealed substantial variation in antigen prevalence by region (**Figure 2**), with the global top-5 K loci estimated to cover 56% of neonatal sepsis isolates in Eastern Africa and 44% in Southern Africa, but only 38% and 28% in South Asia and Western Africa, respectively (**Figure 3**, **Table 2**). This variation was partly driven by differences in geographic distribution of clones carrying distinct K loci (**Appendix S1**). Notably, the global top-20 K loci collectively covered only 67.8% and 50.5% of neonatal sepsis isolates from South Asia and Western Africa, respectively, lower than the 70% threshold used to model the potential impact of a maternal vaccine [10] and proposed as a target in the *K. pneumoniae* vaccine value profile [7]. While the estimated population coverage for any single region could be substantially improved by selecting the top-20 K loci in that region, any region-focused antigen set would provide sub-optimal coverage in other regions (**Figure S4)**. However, the minimum coverage estimate of 70% could be reached for all regions using a single set of 20 K loci, by selecting the eight highest ranking K loci in each region (**Figure 2, Table S4**).

We report primary coverage estimates based on raw counts, rather than cluster-adjusted counts, as it is not anticipated that a maternal vaccine would reduce *K. pneumoniae* colonisation or nosocomial transmission, rather the expected effect is through protection against invasive disease following colonisation of the neonate. Coverage estimates using cluster-adjusted counts are also presented in Tables 2, S4, and S5, and are generally a few percentage points lower than the primary estimates, but are likely much lower than the true expected coverage given the high frequency of nosocomial transmission in LMIC neonatal care settings. We estimated similar cumulative coverage for neonates who died during followup (**Figure 3**, **Table 2**), although we were not able to discern how many of these fatalities were a direct result of *K. pneumoniae* sepsis. We were also unable to assess differences in case fatality rate by K locus or O type, due to sample size limitations and the lack of direct assessment of cause of death. Future estimation of KL- or O-specific fatality rates, and invasiveness (i.e. the probability of causing an infection given colonisation), could provide useful context to prioritise antigen targets, as has been shown for *S. pneumoniae* [51]. While capsule overexpression and hypermucoidy in *K. pneumoniae* are known to promote increased virulence [52,53], very few neonatal sepsis isolates in this study carried the associated *rmpADC* hypermucoidy locus (n=15, 0.8%) [54].

The vast majority of isolates included in our analyses were predicted to produce ESBLs and there was little difference in the cumulative coverage of the top-20 K loci among ESBL producing isolates compared to total isolates. Nor was there a substantial coverage difference for the subset of isolates predicted as carbapenemase producers (**Table 2**). We therefore recommend that genomic surveillance efforts to inform vaccine design should aim to capture the largest possible sample size by including all neonatal sepsis cases regardless of AMR. Subgroup analyses for AMR phenotypes of interest can be conducted post-hoc to check for variation in coverage, as we have shown here.

O types are far less diverse than K types in *K. pneumoniae*, and here we estimated the top-5 O types (two dominant O1 subtypes, plus O2⍺, O2β and O4) collectively covered 86% of total isolates (76–92% per region). Together with the next five most common O types (O3 variants, O5, O13 and O15), we estimate that ∼99% of isolates could be covered in all regions (**Figure 4, Table S5**), i.e. a 10-valent O-based vaccine could theoretically protect against ∼99% of infections. However, there remains uncertainty about the protective efficacy of antibodies targeting the O antigen, due to the possibility of masking by the capsule [16,17,55,56]. There is evidence that immunisation with an O1 containing vaccine does not protect mice against K2/O1 *K. pneumoniae* (irrespective of hypermucoidy) [16,57], which is concerning given that our data showed KL2, encoding K2 biosynthesis, was the most common K locus. Notably, the recent phase III trial of the ExPEC9V anti-O vaccine targeting extraintestinal pathogenic *Escherichia coli* – a close relative of *K. pneumoniae* that produces similar K and O antigens – was aborted because it was found to be insufficiently effective at preventing invasive *E. coli* disease in older adults in high-income countries (clinical trial reference: NCT04899336).

We caution against broad generalisation of our findings to other target populations or disease manifestations, e.g. healthcare-associated infections in adults in low or high resource settings. Our data show that geographic variation in circulating clones (each with dominant K locus associations) results in geographic variation of K locus prevalence, and it is well established that similar geographic clone variation exists among adult disease-causing populations [32,44,58]. While our global top-5 K loci were all reported among the top 10 causes of adult bloodstream infections in Oxford (UK) [59], only KL2 and KL62 were among the top 10 causes of bloodstream infections reported from South and Southeast Asia [32]. Additional comparisons are limited because most studies of adult *K. pneumoniae* infections are skewed towards outbreak clusters and/or CP strains, highlighting a key limitation for the design of vaccines to prevent adult disease.

Our K and O prevalence estimates were derived using a robust Bayesian statistical framework applied to cluster-adjusted antigen counts, to limit the impact of stochastic outbreak clusters. Importantly our sensitivity analyses suggest the findings are generally robust to clustering thresholds and study inclusion in the meta-analysis. Nonetheless, there are a number of limitations that warrant careful consideration.

Firstly, the currently available data have several limitations: i) The total sample size (n=1930) is relatively small given the total possible K loci and O types. ii) Our data captured relatively few countries per region (≤3 countries each, except Eastern Africa with n=6 countries), and with unequal representation per region (**Table 1**). As more data are collected from additional countries we expect that the relative ranks of individual K loci may change. We anticipate that the composition of the overall top-5 will remain fairly stable, because the credible intervals for these loci did not substantially overlap those for lower ranked loci (**Figure 2a**), they were common across all regions (**Figure 2b**) and remained stable in our leave-one-out sensitivity analysis (**Appendix S2**). However the other K loci showed substantial geographic variation, and had broadly overlapping credible intervals for estimated prevalence (**Figure 2a**). As a consequence, the composition of the overall top-20 is likely to change as more countries are considered, especially those from Western Africa which is most underrepresented in the current data. iii) The data were collated from different studies and surveillance programs, each using different protocols and likely influenced by local differences in diagnostic standards, stewardship and prophylactic antimicrobial use.

Together these differences may have contributed to differential ascertainment bias, e.g. the samples at some sites may be relatively skewed towards more severe cases which could influence antigen profiles if a subset of antigens are associated with more severe disease (although there is currently no evidence to suggest this). iv) Only a subset of studies collected outcome data, and those that did focussed on all-cause mortality within a broad time range (up to 60 days of life or 30 days post enrollment), which means we were unable to directly compare antigen profiles among fatal and non-fatal neonatal sepsis cases, nor to adjust for variation in gestation, birthweight, comorbidities or other risk factors. v) Most sites did not have longitudinal data, and there was confounding of country with time period, hence we could not directly explore temporal fluctuations in KL or O type prevalence. However, we note that the available data indicate that regional differences could not be explained by differences in the calendar timing of samples from different regions (**Appendix 1**), and those sites with longitudinal data showed reasonable stability within individual sites over time (**Figure S6**).

Secondly, our analyses have relied upon genomic antigen predictions rather than direct serological data, which is very challenging to obtain. K typing data for 731 isolates, including a subset of those from the BARNARDS, GBS-COP and MLW studies, indicates a high level of concordance between genomic predictions and serological phenotypes, in particular for isolates carrying one of the global top-20 K loci (84.5% concordance overall, 91.4% for top-20 K loci); however, there is still room for improvement [29]. Additionally, only half of the known K loci correspond to serologically defined K types and/or published glycan structures. While the available data indicate that unique loci correspond to unique polysaccharide structures, it remains unclear if these structures are serologically distinct, meaning the genomic predictions may overestimate the true antigen diversity (and thus underestimate vaccine coverage). Conversely, genetic variation within K loci, i.e. allelic variation within copies of the same K locus type, may create differences in antibody binding, meaning the genomic predictions may underestimate antigen diversity (and thus overestimate vaccine coverage). Ongoing work from our team and collaborators aims to better understand the link between K genotype and phenotype (serological, structural, and immunological), to support continued improvements for genomic predictions. This will include the generation of additional serological phenotype data. However, it is not feasible to serotype all isolates because the process is labour intensive and currently only available at a single reference laboratory at the Statens Serum Institut (Denmark), which poses resource constraints and accessibility issues e.g. due to shipping and export permit requirements for Nagoya protocol compliance. Similarly, there is currently no broadly accessible O antigen typing protocol, and although the genetic determinants of O serotypes are considered comparatively very well understood [22], there are no data available with which to perform a formal assessment of the predictive accuracy of our genomic approaches. There is therefore a clear need for low cost and broadly accessible K and O serotyping approaches to support sero-epidemiology analysis, that could inform preliminary vaccine design and ongoing surveillance to monitor for population shifts following vaccine introduction.

Thirdly, our analyses have not attempted to capture information about potentially cross-reactive serotypes. For example, there is converging evidence that antibodies raised against individual O1 or O2 subtypes may be cross-protective against other O1 and O2 subtypes, respectively [16,17]. In that regard, the total number of O antigens required to reach maximal population coverage may be lower than those reported here which are based on individual subtypes. Similarly, there is emerging evidence that antibodies raised against serologically distinct K types may have cross-binding and cross-protective activity [60]. More work is needed to fully assess these cross-reactions and understand their relevance for protection against *K. pneumoniae* disease. However, if cross-protection is confirmed for one or more of the top-ranked K loci, the population coverage we have reported would be considered underestimates.

Finally, while this work has focussed solely on polysaccharides, we acknowledge that protein antigens are likely to be much less variable across the population. However, polysaccharide vaccines are currently more popular vaccine targets, as polyvalent *K. pneumoniae* K vaccines have been shown to be safe and immunogenic in humans [56], and single or bivalent anti-K vaccines can be effective in preventing invasive *K. pneumoniae* infection in murine models [61,62]. While protein-based vaccines are also in development [63], these are unlikely to be suitable for a maternal vaccine to be deployed in low-income countries as they typically require more than one dose to induce protection, and the timeframe for immunisation in pregnancy is limited, especially given that up to half of neonates who develop sepsis are born preterm [3].

Our data suggest that it could be possible to achieve a target 70% population coverage of circulating K serotypes in all key geographic regions with a single 20-valent anti-K vaccine, particularly if there turns out to be cross-reactivity between antigens produced by distinct K loci. Greater coverage would likely be achieved, with lower valency, using locally-targeted formulations. However, due to the complex nature of glycoconjugate production, development of multiple high-valency formulations would generate additional manufacturing and regulatory complexities with important cost and logistical considerations. On the other hand, this scenario may represent an important opportunity to support and expand local manufacturing capabilities and processes, of the kind demonstrated to great effect during the COVID-19 pandemic [64,65]. In either scenario, additional K type data will be required to support antigen prioritisation and ensure adequate population coverage per region, e.g. including a greater number of countries and sites per region, particularly for those regions that were underrepresented in our meta-analysis. Longitudinal sampling will also be essential to understand the extent of temporal fluctuations and the impact on theoretical vaccine coverage. Our preliminary analysis suggests a level of stability sufficient to maintain 20-valent vaccine coverage >70% over a 5–10 year timeframe, which is very encouraging but should be considered with caution until further data are available. An O antigen based vaccine could theoretically provide much greater population coverage than a K based vaccine and at lower valency. However, much more data is needed to understand the potential efficacy of such a vaccine, given growing evidence about capsular masking. A combination K and O vaccine may offer an alternative solution, for example if it is found that some key K antigens are more or less likely than others to mask O antigens.

## Data Availability

All data produced are available in the manuscript or available online at https://github.com/klebgenomics/KlebNNSsero and in interactive form via a Shiny web application at https://klebsiella.shinyapps.io/neonatal/. Whole-genome sequence data were deposited by the originating study teams in INSDC databases, under the following BioProjects: BARNARDS, PRJEB33565; SPINZ, PRJEB46513; MLW, PRJEB42462; NIMBIplus, PRJNA1168993; DH, PRJEB70311; BabyGERMS, PRJNA79648; CHRF, PRJEB90555; NeoOBS-India, PRJEB70311; GBS-COP, PRJNA1175467; Kilifi, PRJNA1265413; MBIRA: PRJNA1274034, NeoBAC, PRJNA1265413; AKU, PRJNA126164.

https://klebsiella.shinyapps.io/neonatal/

## Acknowledgments

We thank the participants and their families in all the contributing studies, and the clinical and laboratory staff involved in collection and processing of relevant samples and isolates. This work was supported by Monash eResearch capabilities, including M3. We would also like to acknowledge the expert support of the sequencing and computational genomics teams at the CHOP Microbiome Core (Philadelphia), CHRF (Dhaka), Infectious Diseases Research Laboratory (Karachi), International Livestock Research Institute (Nairobi), Kilimanjaro Clinical Research Institute (Moshi), Neuberg Center for Genomic Medicine (Ahmedabad), NICD (Johannesburg), Quadram Institute (Norwich), Wellcome Centre for Human Genetics (Oxford), and Wellcome Sanger Institute (Cambridge); and the BARNARDS microbiology and genomics team at Cardiff University.

## Author Contributions

**Conceptualization:** Kathryn E Holt, Kelly L Wyres

**Data Curation:** Jabir A Abdulahi, Anne V Amulele, Matthew Bates, Eva Heinz, Yogesh Hooda, Weiming Hu, Kajal Jain, Samiah Kanwar, Rindidzani Magobo, Courtney P Olwagen, John M Tembo, Tolbert Sonda, Jonathan Strysko, Caroline C Tigoi

**Formal Analysis:** Shaun P Keegan, Thomas D Stanton, Kathryn E Holt, Kelly L Wyres **Funding Acquisition:** Alexander M Aiken, James A Berkley, Susan E Coffin, Nicholas A Feasey, Nelesh P Govender, Davidson H Hamer, Shabir A Madhi, M Imran Nisar, Samir K Saha, Senjuti Saha, M Jeeva Sankar, Ramesh K Agarwal, Kathryn E Holt, Kelly L Wyres **Investigation:** Sameen Ahmad Amin, Kyle Bittinger, Jennifer Cornick, Ebenezer Foster-Nyarko, Wilson Gumbi, Aneeta Hotwani, Naveed Iqbal, Steven M Jones, Furqan Kabir, Waqasuddin Khan, Chileshe L Musyani, Carolyn M McGann, Varsha Mittal, Ahmed M Moustafa, Patrick Musicha, James CL Mwansa, Moreka L Ndumba, Erkison E Odih, Donwilliams O Omuoyo, Oliver Pearse, Laura T Phillips, Paul J Planet, Aniqa Abdul Rasool, Charlene MC Rodrigues, Kirsty Sands, Arif M Tanmoy, Erin Theiller, Allan M Zuza **Methodology:** Shaun P Keegan, Thomas D Stanton, Kathryn E Holt, Kelly L Wyres **Resources:** Sulagna Basu, Grace J Chan, Kenneth C Iregbu, Jean-Baptiste Mazarati, Semaria Solomon Alemayehu, Timothy R Walsh, Rabaab Zahra, Kajal Jain, Angela Dramowski, Sombo Fwoloshi, Appiah-Korang Labi, Lola Madrid, Noah Obeng-Nkrumah, David Ojok, Boaz D Wadugu, Andrew C Whitelaw, Adhisivam Bethou, Anudita Bhargava, Atul Jindal, Ruchi N Nanavati, Priyanka S Prasad, Apurba Sastry, Joveria Q Farooqi, Najia Ghanchi, Fyezah Jehan, Erum Khan, Alexander M Aiken, James A Berkley, Susan E Coffin, Nicholas A Feasey, Nelesh P Govender, Davidson H Hamer, Shabir A Madhi, M Imran Nisar, Samir K Saha, Senjuti Saha, M Jeeva Sankar, Ramesh K Agarwal

**Software:** Shaun P Keegan, Thomas D Stanton, Kathryn E Holt, Kelly L Wyres **Supervision:** Sulagna Basu, Grace J Chan, Kenneth C Iregbu, Jean-Baptiste Mazarati, Semaria Solomon Alemayehu, Timothy Walsh, Rabaab Zahra, Kajal Jain, Angela Dramowski, Sombo Fwoloshi, Appiah-Korang Labi, Lola Madrid, Noah Obeng-Nkrumah, David Ojok, Boaz D Wadugu, Andrew C Whitelaw, Adhisivam Bethou, Anudita Bhargava, Atul Jindal, Ruchi N Nanavati, Priyanka S Prasad, Apurba Sastry, Joveria Q Farooqi, Najia Ghanchi, Fyezah Jehan, Erum Khan, Alexander M Aiken, James A Berkley, Susan E Coffin, Nicholas A Feasey, Nelesh P Govender, Davidson H Hamer, Shabir A Madhi, M Imran Nisar, Samir K Saha, Senjuti Saha, M Jeeva Sankar, Ramesh K Agarwal

**Visualization:** Shaun P Keegan, Thomas D Stanton, Kathryn E Holt, Kelly L Wyres

**Writing – Original Draft Preparation:** Kathryn E Holt, Kelly L Wyres

**Writing – Review & Editing:** All authors.

## Financial Disclosure Statement

This study was supported by the Gates Foundation [https://www.gatesfoundation.org/] (grants INV049364, INV025280, INV077266 to KEH; INV049641 to KLW; INV003519 to AAiken; INV005180 to NAF; INV041685 to JAB; INV008112 to NPG; INV005567 to CT; INV005691 to DHH; INV023821 to Senjutia Saha, INV005773 to SAM, INV046521 and INV080845 to MIN, INV065400 to AMM, INV065400 to SC), the Fleming Fund [https://www.flemingfund.org/] (grant FF25-286 for SeqAfrica to support sequencing capacity at KCRI), the Center for AIDS Research [https://www.niaid.nih.gov/research/centers-aids-research] (core support grant to SC), the Thrasher Research Fund [https://www.thrasherresearch.org/] (grant #12036 to DHH), the Wellcome Trust [https://wellcome.org/] (grant 217303/Z/19/Z to EH, core support grants 206194 to WSI, 206454 to MLW, 203077/Z/16/Z to KWTRP), and the National Health and Medical Research Council of Australia [https://www.nhmrc.gov.au/] (APP1176192 to KLW). Pfizer [https://www.pfizer.com/] sponsored the sequencing of a subset of isolates from the GBS-COP study for WITS-Vida (SAM). The funders had no role in study design, data collection and analysis, decision to publish, or preparation of the manuscript. The conclusions and opinions expressed in this work are those of the author(s) alone and shall not be attributed to any funder. Under the grant conditions of the Gates Foundation and Wellcome Trust, a Creative Commons Attribution 4.0 License has already been assigned to the Author Accepted Manuscript version that might arise from this submission. Please note works submitted as a preprint have not undergone a peer review process.

## Notes

### Competing Interest Statement

The authors have declared no competing interest.

### Author Declarations

The Observational/Interventions Research Ethics Committee of the London School of Hygiene and Tropical Medicine gave ethical approval for this work.

### Summary of Updates

The model specification and explanation in the Methods section has been updated for clarity. Minor updates were made to the Discussion, primarily to note that coverage estimates using cluster-adjusted counts are also presented (in Tables 2, S4, and S5), but are likely much lower than the true expected coverage given the high frequency of nosocomial transmission in LMIC neonatal care settings. Details of the source of the base map underlying Figure 1 were updated.

