## Supporting Information and Appendices for "Distribution of capsule and O types in *Klebsiella pneumoniae* causing neonatal sepsis in Africa and South Asia: meta-analysis of genome-predicted serotype prevalence to inform potential vaccine coverage"

#### Code and data availability

All data and code used to generate the material presented here are available from: github.com/klebgenomics/KlebNNSsero (DOI: 10.5281/zenodo.15756796).
An interactive web application, implemented in R shiny, is also available to (i) reproduce and explore the modelled estimates for prevalence and coverage data included in this paper, and (ii) undertake additional analyses of the raw data (e.g. to explore crude pooled estimates of prevalence and coverage for different subsets of loci and/or different subsets of samples). The web application is available at https://klebsiella.shinyapps.io/neonatal/.

### Contents


Supporting Tables

- Table S1. Ethics committees approving individual studies
- Table S2. Studies included in the meta-analysis.
- Table S3. Isolates and sequence data included in the study
- Table S4. Coverage estimates for KL set 1 (global top 20)
- Table S5. Coverage estimates for global top-10 O genotypes

Supporting Figures

- Figure S1. Flow diagram for inclusion of isolates
- Figure S2. Modelled estimates of cluster-adjusted global and regional prevalence of all K loci.
- Figure S3. Posterior distributions of K locus prevalence per region.
- Figure S4. Regional coverage estimates for top 20 K loci selected on a region-focused basis.
- Figure S5. Posterior distributions of O type prevalence per region.
- Figure S6. Longitudinal data.

Appendix S1. Genomic diversity.

- Appendix S1.1: Distribution of K loci and O types by country.
- Appendix S1.2: Distribution of clones per year per site.
- Appendix S1.3: Crude cluster-adjusted annual frequency per K locus per region.
- Appendix S1.4: Crude cluster-adjusted annual frequency per O type per region.
- Appendix S1.5: Region distribution of K and ST combinations associated with O types that demonstrate regional variation.
- Appendix S1.6: Definition of genetically inferred O types used in the analysis

Appendix S2. Sensitivity analyses.

- Appendix S2.1 Sensitivity of K locus global ranks to study inclusion.
- Appendix S2.2 Sensitivity of O type global ranks to study inclusion.
- Appendix S2.3 Effect of cluster adjustment on crude proportions per site.
- Appendix S2.4 Effect of cluster adjustment on Bayesian modelled estimates of global K locus prevalence.
- Appendix S2.5 Sensitivity analysis showing effect of temporal clustering thresholds on K locus proportions and ranks (modelled global estimates).
- Appendix S2.6 Sensitivity analysis showing effect of temporal clustering thresholds on K locus proportions and ranks (crude proportions).
- Appendix S2.7 Sensitivity analysis showing effect of SNP clustering thresholds on K locus proportions and ranks (crude proportions).
- Appendix S2.8: Coverage estimates for KL set 1 (global top 20)
- Appendix S2.9: Coverage estimates for KL set 2 (top-8 per region)
- Appendix S2.10: Coverage estimates for O types

Appendix S3. Model Diagnostics.

- Model diagnostics for all Bayesian models

#### Supporting Tables

##### Table S1. Ethics committees approving individual studies

Approval for the meta-analysis presented here was granted by the Observational / Interventions Research Ethics Committee of the London School of Hygiene and Tropical Medicine (ref #29931), and covers inclusion of data from the studies whose primary ethical approvals are listed in this table.

| Study | Ethics Committees granting approval |
|---|---|
| **Baby GERMS-SA** | Human Research Ethics Committee of the University of the Witwatersrand (M190320). Approvals for the tier 2 surveillance study were received from each provincial research committee through registration on the National Health Research Database. |
| **BARNARDS** | Ethical Review Committee, Bangladesh Institute of Child Health, BICH-ERC-4/3/2015, 15/09/2015. Boston Children's Hospital, IRB-P00023058, 11/08/2016. Institutional Ethics Committee, National Institute of Cholera and Enteric Diseases and Institute of Post Graduate Medical Education and Research, A-I/2016-IEC, 17/11/2016. IPGMER Research Oversight Committee, Inst/IEC/2016/508, 04/11/2016. Kano State Hospitals Management Board, 8/10/1437AH, 13/07/2016. Health Research Ethics Committee (HREC), National Hospital, Abuja, NHA/EC/017/2015, 27/04/2015. Republic of Rwanda National Ethics Committee, No342/RNEC/2015, 10/11/2015. Stellenbosch University and Tygerberg Hospital, Research projects, Western Cape Government, N15/07/063, 04/12/2015 and 02/02/2016. |
| **KWTRP surveillance** | KEMRI Scientific and Ethics Review Unit, ref 281/4687, 17/4/2023. A research license was obtained from the National Commission for Science, Technology and Innovation, ref 527823, license no. NACOSTI/P/23/26005, 31/5/2023. |
| **GBS-COP** | University of the Witwatersrand, Human Research Ethics Committee (HREC), ref 181110, 24/4/2023. |
| **MLW Biobank** | University of Malawi College of Medicine Research Ethics Committee (COMREC) (P.11/18/2541). |
| **SPINZ** | Boston University Medical Center Institutional Review Board, USA (ref H-33473), 21/3/2016. Excellence in Research Ethics and Science (ERES) CONVERGE, Zambia (ref 2015-Jan-004), 1/3/2015. |
| **NIMBIplus** | NIMBI study: University of Pennsylvania IRB (ref 833786), 22/4/2020. Children’s Hospital of Philadelphia Research Institute IRB (ref 19-016848), 25/7/2020. IRBs of Princess Marina Hospital IRB (PMH 2/11AII(372)), 20/3/2024 and the Health Research Development Committee (HRDC) in Botswana (ref HPRD 6/14/1), 15/2/2024. |

| | |
|---|---|
| | SHARE study: University of Pennsylvania IRB (ref 851492), 9/6/2020. Princess Marina Hospital (ref PMH 2/2A(7)/201), 20/5/2022. University of Botswana IRB (ref UBR/RES/IRB/BIO/205), 11/5/2022. Health Research & Development Committee (HRDC) in Botswana (HPDME:13/18/1), 21/4/2022. |
| **MBIRA** | London School of Hygiene and Tropical Medicine HREC, ref 21236-4, latest amendment approved 27/3/2024. Korle Bu Teaching Hospital Institutional Review Board, 00097/2020, 18/11/2020; National Health Research Authority, 17/2/2020; Ministry of Science and Higher Education, 1.16/10.81/13, 24/2/2021; Western Cape Government, N20/02/072; National Institute for Medical Research, R.8a/Vol.IX/3575, 11/12/2020; Combined Research and Ethics Committee Swarm Hub research protocol v2, 2/11/2020. |
| **AKU** | Aga Khan University Ethics Review Committee, ref 2023-8485-25083, last renewed 13/5/2024. |
| **AIIMS** | Institute Ethics Committee, All India Institute of Medical Sciences, New Delhi, ref IEC-683/07.12.2018, RP-12/2018, 18/12/2018. |
| **CHRF** | Bangladesh Institute of Child Health HREC, ref BCH-ERC-02-02-2021, 3/2/2021. |

#### Table S2. Studies included in the meta-analysis.

Details of type of study, number of isolates; laboratory methods for identification, storage, DNA extraction, sequencing; informatics methods for quality control and assembly; BioProject accession and citation/s for primary study.

[see TSV file:
https://github.com/klebgenomics/KlebNNSsero/blob/main/tables/TableS2_StudyMethodsInfo.tsv]

#### Table S3. Isolates and sequence data included in the study

[see TSV file:
https://github.com/klebgenomics/KlebNNSsero/blob/main/tables/TableS3_sampleInfo.tsv]

#### Table S4. Coverage estimates for KL set 1 (global top 20)

Cumulative coverage estimates for the top 5, 10, 15 or 20 K loci (KL) ordered by cluster-adjusted global prevalence estimate. For each geographic location, the breakdown of isolate counts and corresponding coverage estimates are shown for subgroups defined by types of isolates (ESBL, extended-spectrum beta-lactamase; CP, carbapenemase-producing) or outcome of infection (fatal infections only). Coverage estimated from raw and cluster-adjusted counts ('adj') are shown.

| | Counts | N | 5 KL | 10 KL | 15 KL | 20 KL |
|---|---|---|---|---|---|---|
| **Global** | raw | 1930 | 48.8 (46.0-51.7) | 58.7 (55.6-61.9) | 67.0 (63.6-70.4) | 72.8 (69.3-76.4) |
| | adj | 805 | 36.3 (32.4-40.3) | 49.8 (45.3-54.6) | 60.2 (55.2-65.4) | 69.0 (63.6-74.6) |
| Fatal infections | raw | 394 | 49.2 (43.1-55.7) | 58.8 (51.8-66.1) | 70.0 (62.3-77.9) | 75.8 (67.8-84.2) |
| | adj | 214 | 40.1 (31.5-49.7) | 50.3 (40.5-61.2) | 63.7 (52.5-76.3) | 74.5 (62.3-88.1) |
| ESBL infections | raw | 1747 | 50.8 (47.8-53.9) | 60.8 (57.5-64.2) | 69.5 (65.9-73.1) | 75.0 (71.2-78.8) |
| | adj | 683 | 38.2 (33.9-42.7) | 52.3 (47.3-57.6) | 63.5 (57.9-69.4) | 72.1 (66.1-78.3) |
| CP infections | raw | 525 | 40.9 (35.8-46.2) | 51.3 (45.5-57.4) | 65.7 (59.1-72.6) | 71.7 (64.7-78.9) |
| | adj | 228 | 24.0 (18.0-30.7) | 41.5 (33.6-50.3) | 56.6 (47.1-66.8) | 68.5 (58.0-79.8) |
| **Eastern Africa** | raw | 1119 | 55.9 (52.0-60.0) | 65.8 (61.5-70.3) | 70.7 (66.2-75.4) | 75.7 (71.1-80.6) |
| | adj | 367 | 42.1 (36.2-48.6) | 55.5 (48.5-62.9) | 63.8 (56.3-71.7) | 71.0 (63.1-79.4) |
| Fatal infections | raw | 237 | 56.4 (48.7-64.6) | 64.5 (55.9-73.6) | 70.2 (60.9-79.8) | 74.2 (64.8-84.2) |
| | adj | 100 | 45.5 (32.9-59.8) | 54.4 (40.5-69.9) | 64.9 (49.7-82.2) | 73.6 (57.2-92.0) |
| ESBL infections | raw | 1019 | 57.1 (52.9-61.3) | 67.3 (62.7-72.0) | 72.2 (67.4-77.0) | 77.0 (72.1-82.0) |
| | adj | 306 | 45.0 (38.2-52.2) | 59.2 (51.2-67.7) | 67.7 (59.2-76.7) | 74.3 (65.4-83.7) |
| CP infections | raw | 10 | NA | NA | NA | NA |
| | adj | 5 | NA | NA | NA | NA |
| **Southern Africa** | raw | 244 | 43.9 (36.5-51.9) | 50.5 (42.4-59.0) | 71.6 (62.3-81.6) | 76.4 (66.6-86.8) |
| | adj | 151 | 48.7 (39.1-59.4) | 58.1 (47.6-69.5) | 71.3 (59.8-84.3) | 77.7 (65.4-91.4) |
| Fatal infections | raw | 65 | 51.3 (31.9-74.1) | 52.9 (33.1-75.9) | 89.5 (64.4-100.0) | 95.1 (69.1-100.0) |
| | adj | 46 | 55.8 (31.4-85.8) | 60.2 (34.8-90.7) | 77.8 (48.0-100.0) | 87.4 (55.1-100.0) |
| ESBL infections | raw | 191 | 51.9 (42.6-61.8) | 54.6 (45.0-64.7) | 82.1 (70.3-94.1) | 85.2 (73.1-97.6) |
| | adj | 117 | 57.3 (44.7-71.0) | 62.1 (48.9-76.7) | 78.1 (62.9-94.1) | 82.5 (66.9-99.3) |
| CP infections | raw | 66 | 47.8 (30.9-67.4) | 48.0 (31.1-67.6) | 96.7 (74.2-100.0) | 96.9 (74.4-100.0) |
| | adj | 33 | 68.8 (38.4-100.0) | 69.4 (38.8-100.0) | 91.1 (55.7-100.0) | 91.6 (56.1-100.0) |
| **Western Africa** | raw | 76 | 28.4 (18.5-40.3) | 38.2 (26.6-51.7) | 44.5 (31.6-59.3) | 50.5 (36.9-66.4) |
| | adj | 55 | 29.8 (18.4-44.1) | 37.8 (24.7-53.5) | 46.6 (31.8-64.3) | 54.9 (38.6-74.3) |
| Fatal infections | raw | 36 | 19.6 (7.9-36.5) | 40.4 (22.3-62.6) | 51.0 (30.0-75.7) | 60.5 (37.3-87.6) |
| | adj | 28 | 27.8 (12.0-48.5) | 38.5 (19.3-62.8) | 52.1 (29.3-80.4) | 64.4 (37.8-96.4) |
| ESBL infections | raw | 62 | 32.8 (20.2-47.9) | 44.8 (29.5-62.6) | 52.2 (35.5-71.3) | 59.0 (41.1-79.3) |
| | adj | 43 | 34.9 (19.7-54.0) | 43.5 (26.6-64.6) | 54.1 (34.8-77.6) | 63.4 (42.3-88.9) |
| CP infections | raw | 30 | 5.4 (0.2-18.0) | 5.7 (0.3-18.7) | 6.3 (0.4-19.9) | 6.7 (0.4-20.6) |
| | adj | 17 | 9.4 (0.4-30.5) | 10.3 (0.6-32.2) | 11.5 (0.8-34.4) | 12.2 (0.9-35.9) |
| **South Asia** | raw | 491 | 38.3 (33.5-43.6) | 49.9 (44.3-55.8) | 59.6 (53.4-66.4) | 67.8 (61.1-75.0) |
| | adj | 232 | 20.8 (15.7-26.5) | 38.5 (31.5-46.3) | 50.5 (42.4-59.3) | 63.6 (54.6-73.7) |
| Fatal infections | raw | 56 | 35.8 (22.5-51.1) | 50.7 (34.7-69.4) | 66.8 (47.7-88.8) | 78.1 (57.3-100.0) |
| | adj | 40 | 25.7 (12.5-43.1) | 42.6 (24.8-64.3) | 59.7 (38.1-86.3) | 75.2 (50.1-100.0) |
| ESBL infections | raw | 475 | 39.0 (34.1-44.1) | 51.0 (45.2-56.9) | 61.1 (54.8-67.8) | 68.8 (62.0-76.0) |
| | adj | 217 | 20.7 (15.3-26.7) | 39.8 (32.4-47.9) | 52.7 (43.9-62.1) | 65.8 (55.8-76.4) |
| CP infections | raw | 419 | 41.9 (36.4-47.7) | 54.0 (47.6-60.8) | 65.4 (58.2-72.9) | 72.3 (64.7-80.2) |
| | adj | 173 | 20.3 (14.3-27.3) | 40.8 (32.1-50.4) | 56.1 (45.8-67.3) | 70.1 (58.4-82.6) |

#### Table S5. Coverage estimates for global top-10 O genotypes

Cumulative coverage estimates for the 10 O genotypes, ordered by cluster-adjusted global prevalence estimate. For each geographic location, the breakdown of isolate counts and corresponding coverage estimates are shown for subgroups defined by types of isolates (ESBL, extended-spectrum beta-lactamase; CP, carbapenemase-producing) or outcome of infection (fatal infections only).

[see TSV file: https://github.com/klebgenomics/KlebNNSsero/blob/main/tables/TableS5_coverage_O.tsv]

### Supporting Figures

#### Figure S1. Flow diagram for inclusion of isolates

| | AKU | BARNARDS | Baby GERMS | CHRF | DH | GBS-COP | Kilifi | MBIRA | MLW | NIMBIplus | NeoBAC | NeoOBS India | SPINZ |
|---|---|---|---|---|---|---|---|---|---|---|---|---|---|
| **Stored as *K. pneumoniae*** | **130** | **235** | **185** | **472** | **51** | **217** | **201** | **288** | **573** | **40** | **66** | **91** | **432** |
| Not recoverable | → 0 | → 0 | → 4 | → 0 | → 3 | → 112 | → 2 | → 34 | → 111 | → 0 | → 0 | → 0 | → 0 |
| **Subculture & WGS** | **130** | **235** | **181** | **472** | **48** | **105** | **199** | **254** | **462** | **40** | **66** | **91** | **432** |
| Not *K. pneumoniae* | → 1 | → 0 | → 3 | → 0 | → 5 | → 7 | → 14 | → 40 | → 13 | → 2 | → 0 | → 1 | → 78 |
| Fail WGS QC | → 9 | → 0 | → 5 | → 35 | → 1 | → 19 | → 1 | → 41 | → 11 | → 5 | → 0 | → 0 | → 66 |
| **High quality WGS** | **120** | **235** | **173** | **437** | **42** | **79** | **184** | **173** | **438** | **33** | **66** | **90** | **288** |
| Missing year, age, or site | → 47 | → 0 | → 30 | → 0 | → 1 | → 0 | → 0 | → 0 | → 15 | → 0 | → 0 | → 0 | → 2 |
| Pre-2013 | → 0 | → 0 | → 0 | → 146 | → 0 | → 0 | → 16 | → 0 | → 61 | → 0 | → 0 | → 0 | → 0 |
| Duplicate sample | → 0 | → 0 | → 0 | → 0 | → 0 | → 7 | → 0 | → 1 | → 0 | → 4 | → 0 | → 17 | → 0 |
| **Passes metadata filter** | **73** | **235** | **143** | **291** | **41** | **72** | **168** | **172** | **362** | **29** | **66** | **73** | **286** |
| Sites with N<10 pass filters | → 16 | → 17 | → 0 | → 2 | → 7 | → 22 | → 0 | → 8 | → 0 | → 0 | → 0 | → 9 | → 0 |
| **Included in meta-analysis** | **57** | **218** | **143** | **289** | **34** | **50** | **168** | **164** | **362** | **29** | **66** | **64** | **286** |

#### Figure S2. Modelled estimates of cluster-adjusted global and regional prevalence of all K loci.

**(a)** Posterior density distribution for global (overall) prevalence estimates, modelled using cluster-adjusted counts per site, ordered by the mean point estimate. Horizontal lines indicate groups of 10 loci. **(b)** Regional prevalence estimates. Cells are coloured and labelled to indicate the cluster-adjusted regional mean prevalence estimates for each K locus in each region, according to the inset legend.

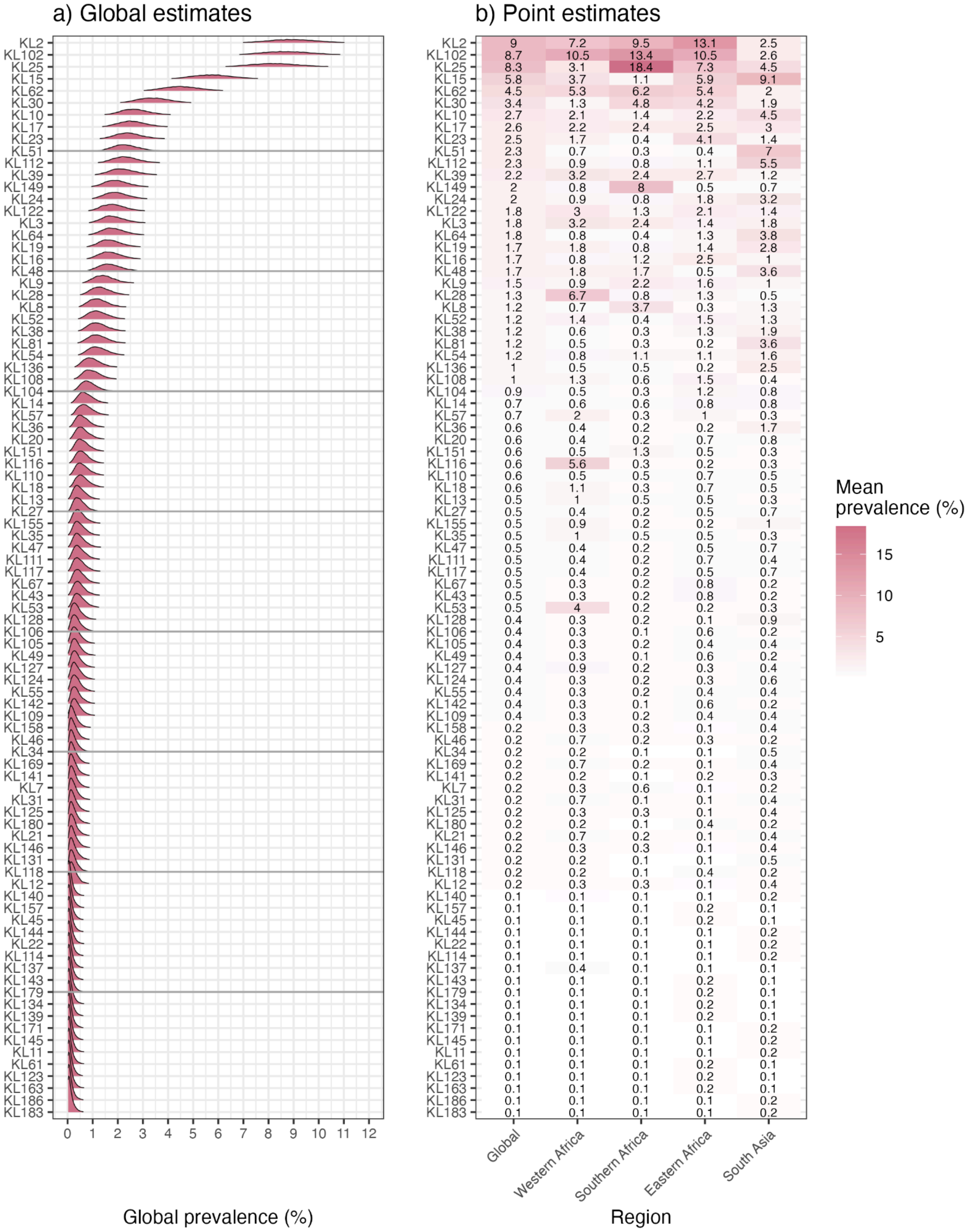

**Figure S3. Posterior distributions of K locus prevalence per region.**

*Indicates K loci for which the median estimate per region differs by >2%, for at least one pair of regions. ^Indicates K loci for which region was a significant linear predictor in a logistic regression model. Horizontal lines indicate the top 10 and 20 global ranked loci.

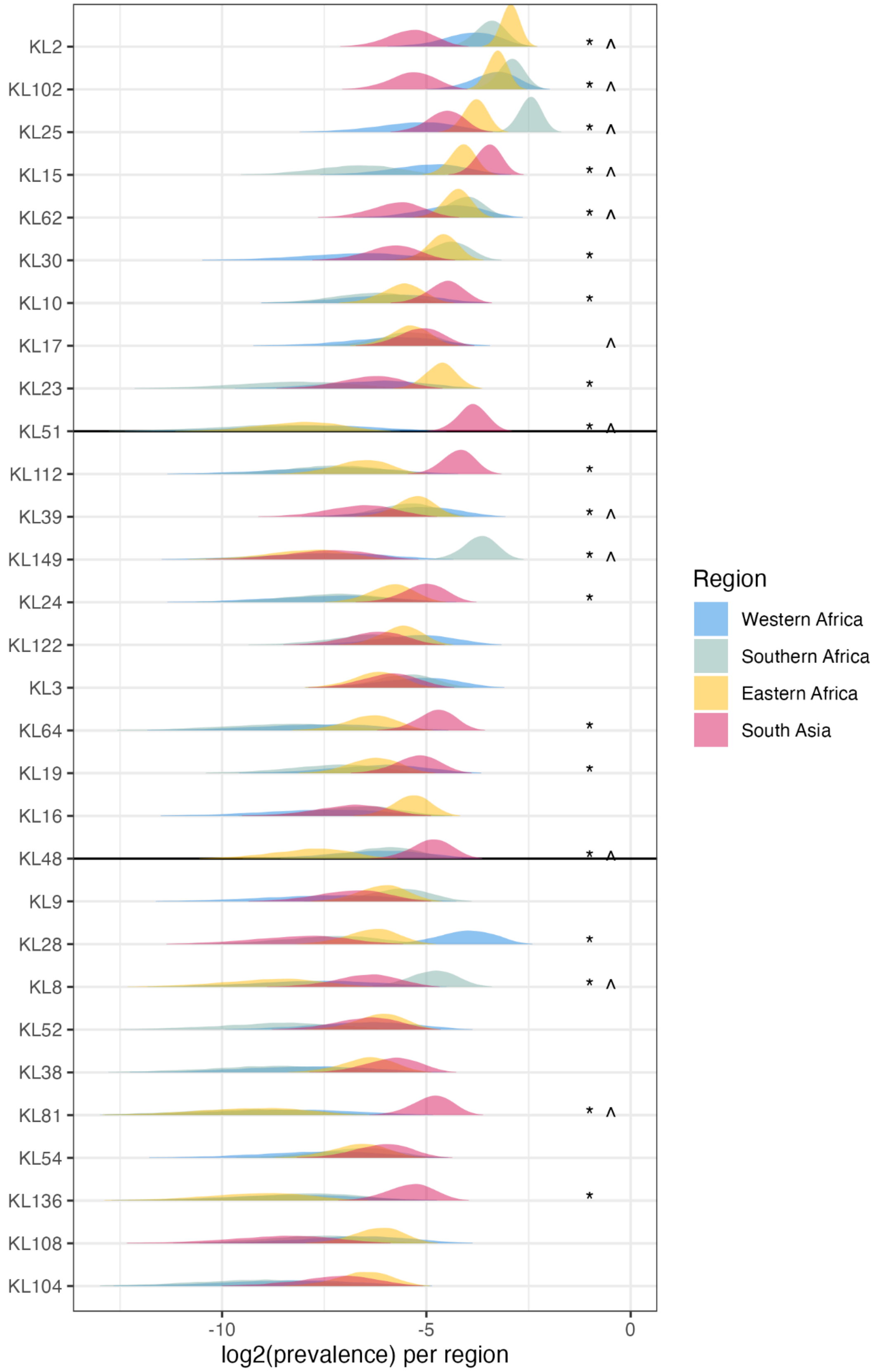

**Figure S4. Regional coverage estimates for top 20 K loci selected on a region-focused basis.**

Each plot shows the modelled coverage in each region, using K loci ranked by cluster-adjusted counts in one region (indicated in the title of the plot).

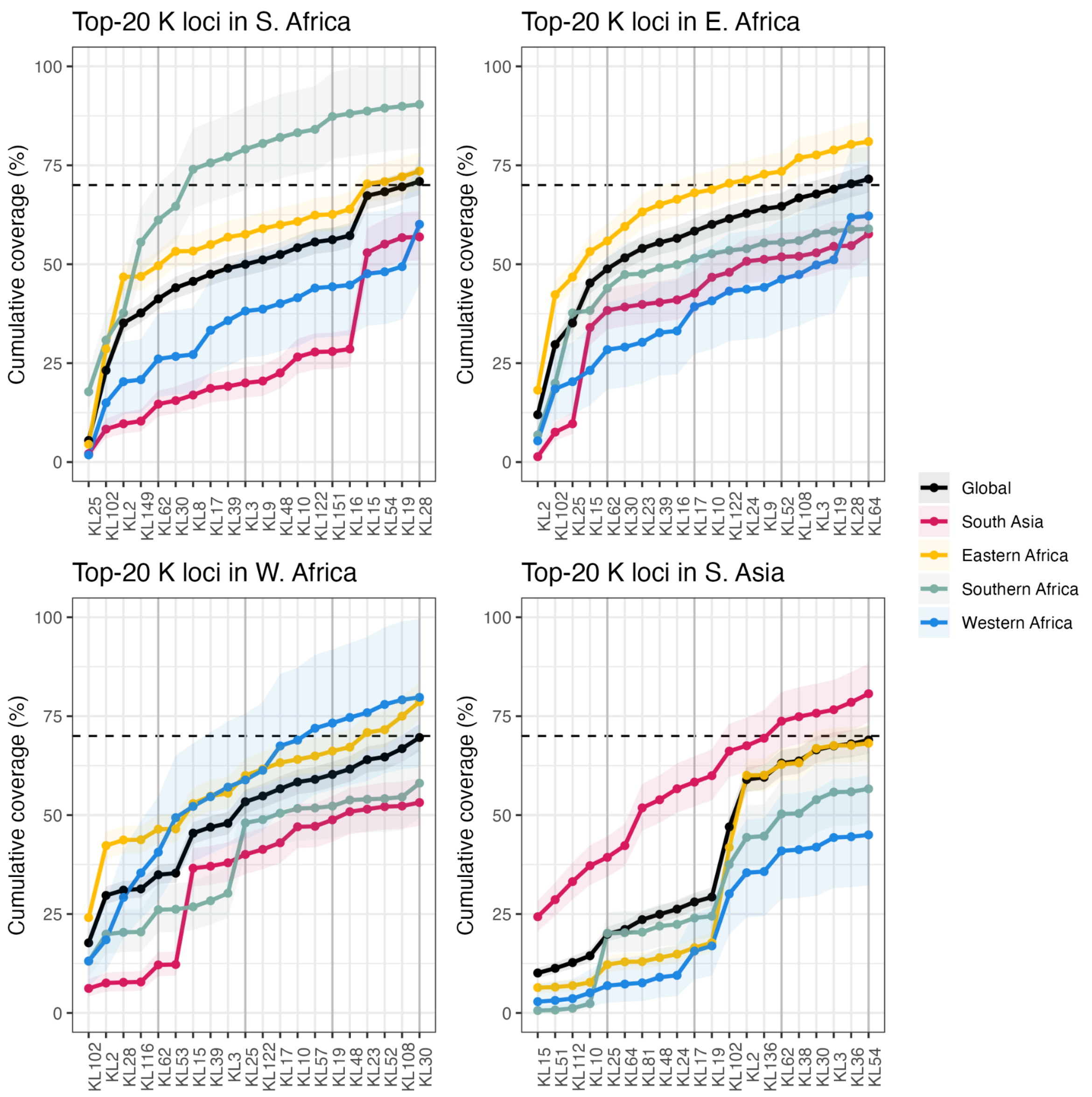

**Figure S5. Posterior distributions of O type prevalence per region.**

*Indicates O types for which the median estimate per region differs by >5%.
^Indicates O types for which region was a significant linear predictor in a logistic regression model.

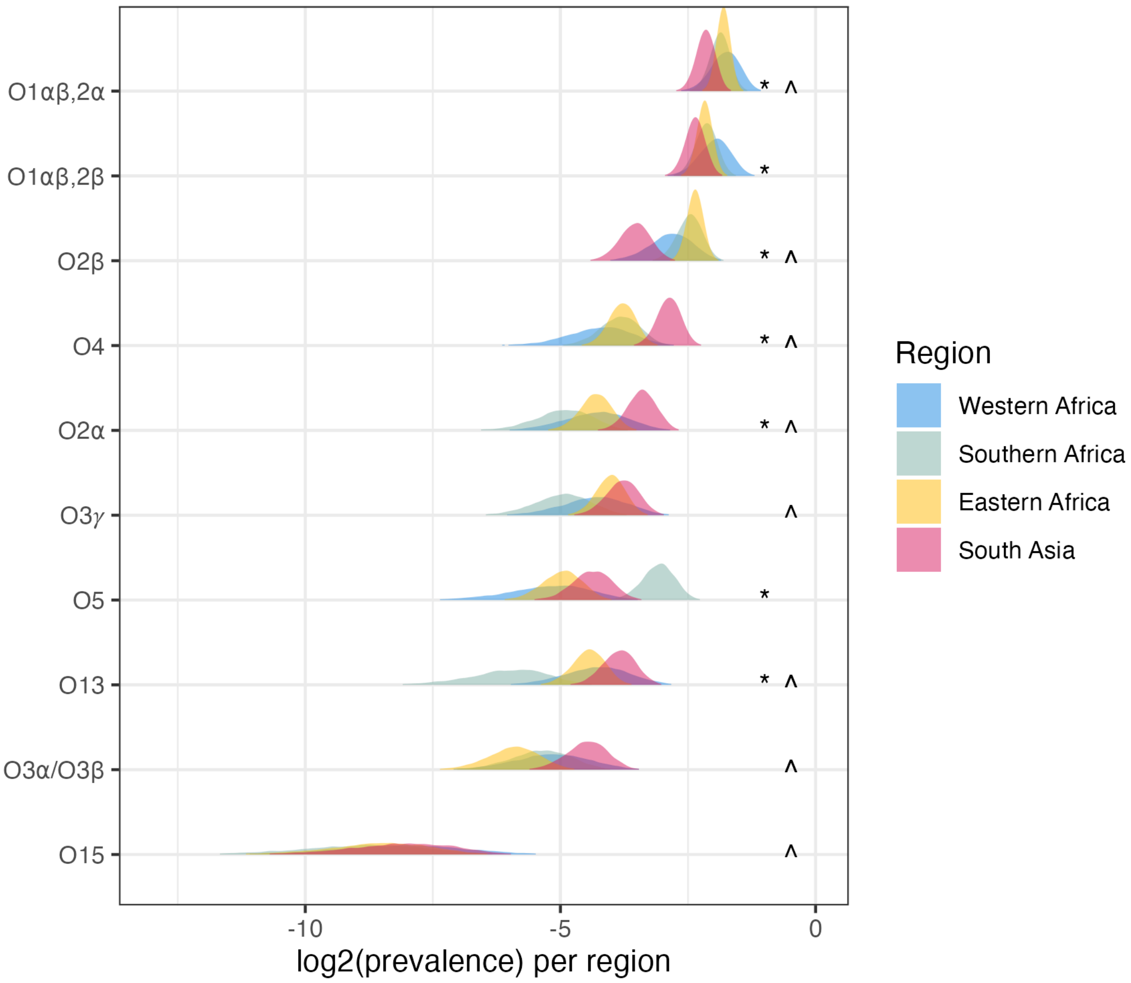

#### Figure S6. Longitudinal data.

Three sites collected routine blood culture isolates for extended periods, providing an opportunity to assess whether a set of loci selected based on prevalence during one surveillance period (here, n=512 samples obtained up to 2017 inclusive, labelled “Training”) could provide coverage of infections occurring in a later time period (here, n=512 data after 2017, labelled “Post-training”). Note the coverage estimates shown reflect the crude proportion per surveillance site (or pooled across sites, labelled “All”), as opposed to Bayesian modelled estimates.

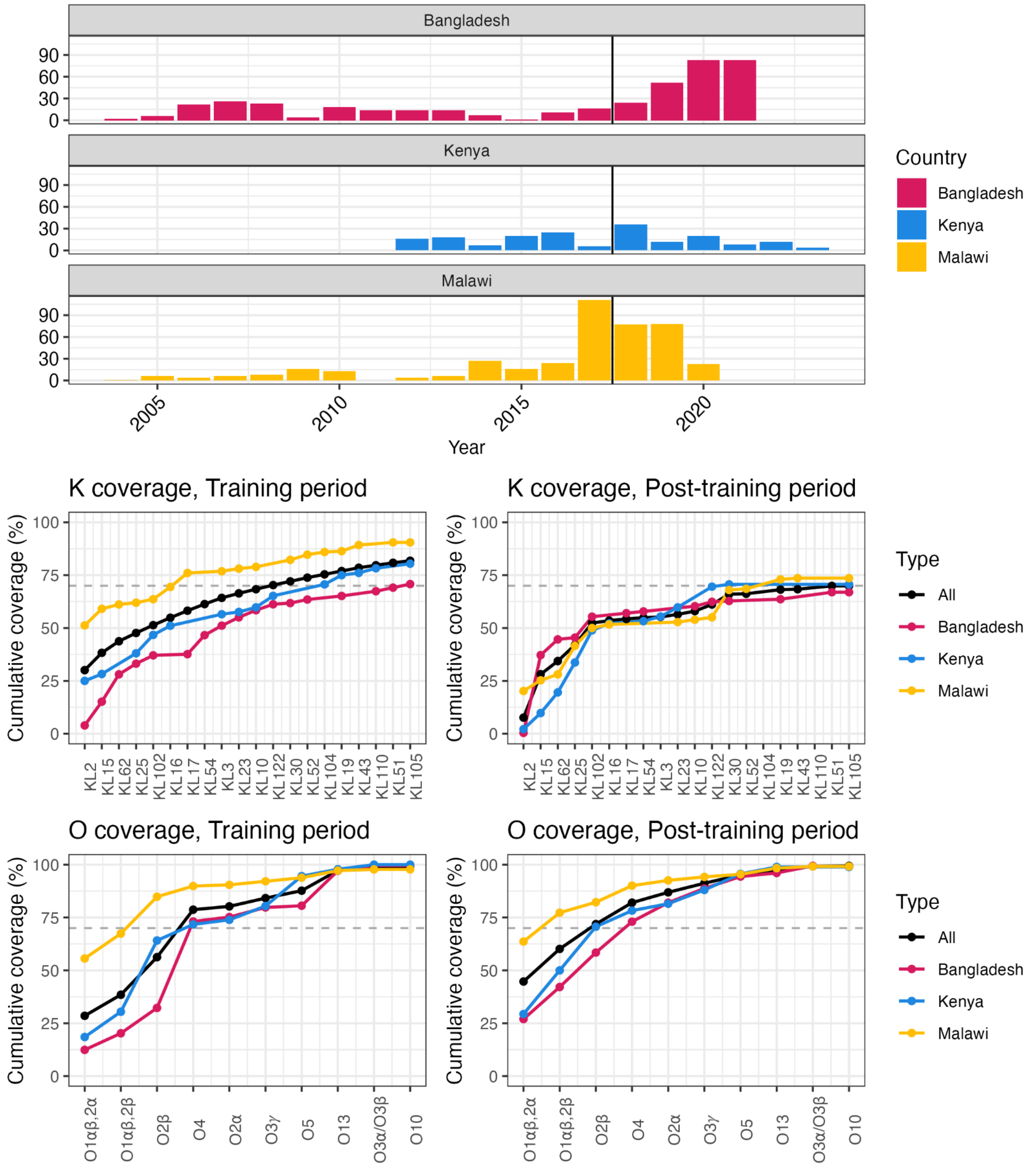

### Appendix S1. Genomic diversity.

#### Appendix S1.1: Distribution of K loci and O types by country.

**(a)** Crude prevalence of K loci across the combined data set (n=1930 high quality genomes from 35 sites in 13 countries). All K loci identified in at least four isolates are included in the plot (n=53/87). Stacked barplot to the left shows the breakdown of common STs (accounting for >10 isolates each) amongst these K loci (coloured as per legend). **(b)** Distribution of K loci by country. Each point represents a single isolate, positioned in the grid to indicate the K locus (row) and country of origin (column), and coloured to indicate the O type (as per legend). Points are jittered around the x and y coordinates so that overlapping points (i.e. genomes of the same K locus and country) are visible.

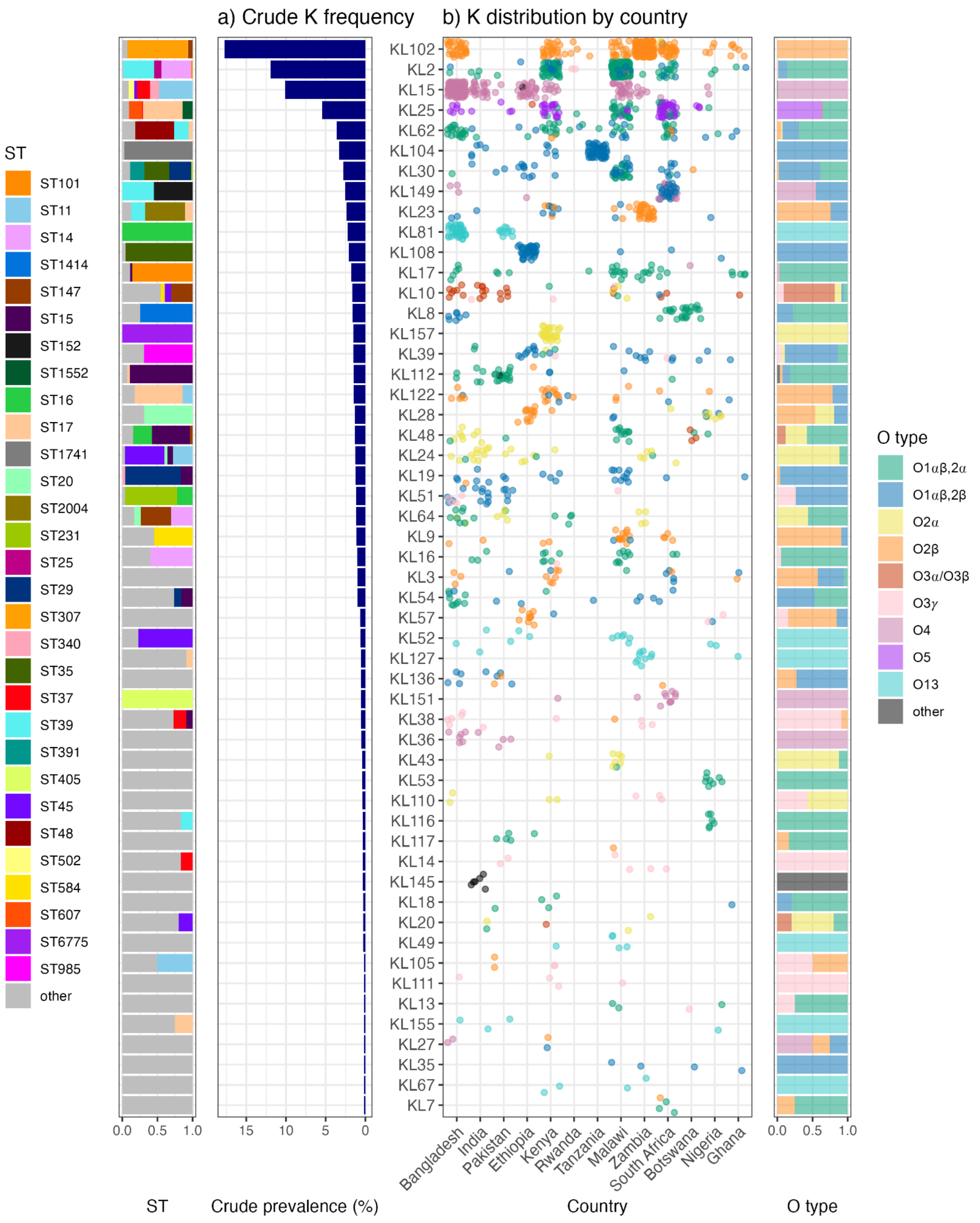

#### Appendix S1.2: Distribution of clones per year per site.

Each panel shows the frequency, per year, of clones (defined as unique combinations of ST/K/O) detected at each study site (arrayed by region and country). Clones that were identified more than 15 times are coloured individually; the rest are grouped as ‘other’.

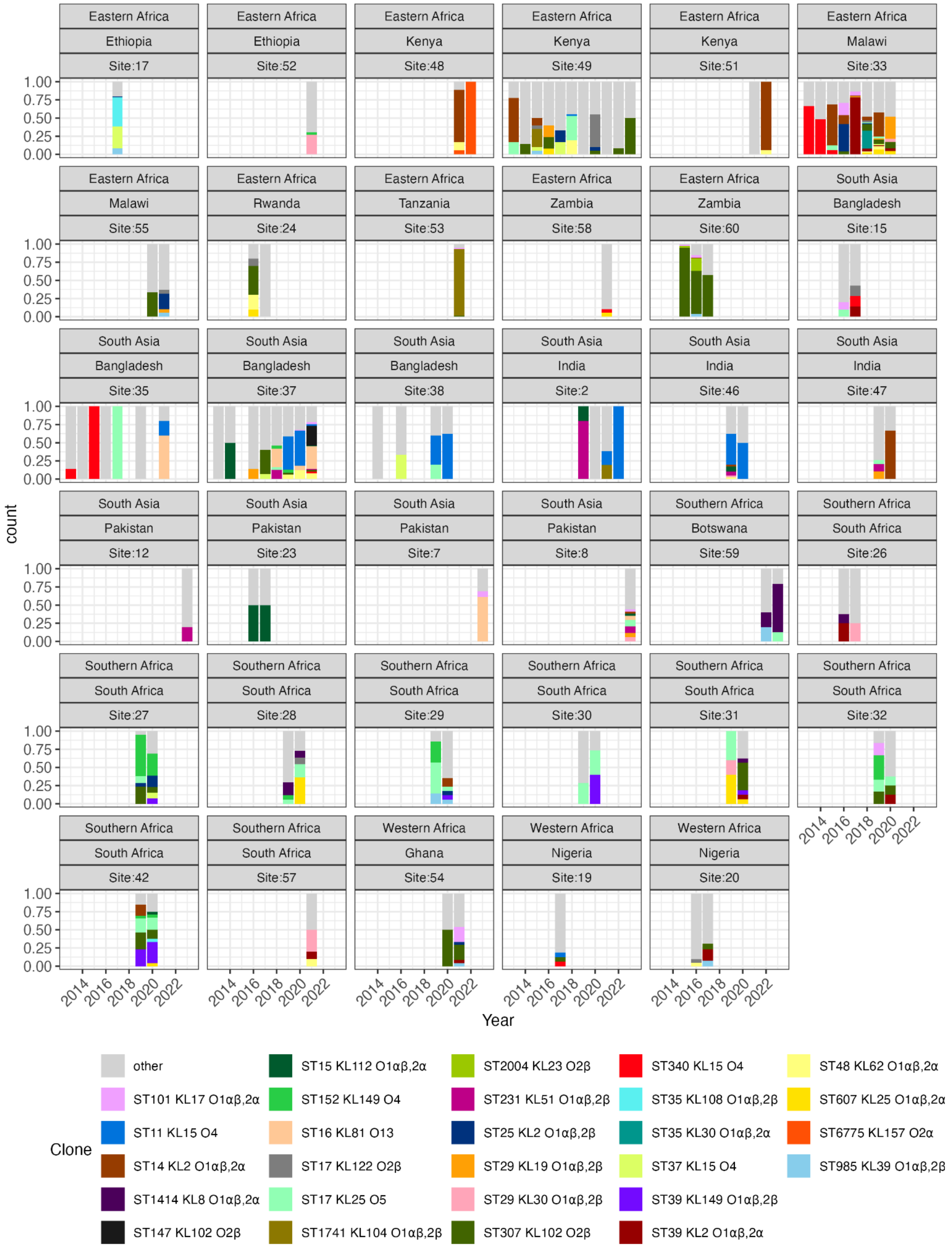

#### Appendix S1.3: Crude cluster-adjusted annual frequency per K locus per region.

Bars show the crude frequency of each K locus per region, using cluster-adjusted counts. Regions are coloured by inset legend. These estimates are pooled across all available studies for that region and year, i.e. the total cluster-adjusted count of isolates per K locus per year in a given region, divided by the total number of unique clusters per year in that region. Plots are labelled with the K locus, and the rank of that K locus according to global Bayesian cluster-adjusted prevalence estimates. Regional variation highlighted in the text cannot be explained by differences in timing; e.g. KL51 were found consistently in Southern Asian isolates between 2018-2023, across 6 sites from 4 studies; but was not detected at all amongst 464 samples from Eastern Africa, 232 from Southern Africa and 26 from Western Africa in this period (KL81 showed similar). In Southern Africa, KL149 was detected at high prevalence in 2019 and 2020 (across 7 sites from three studies), but during this period was detected just twice amongst n=206 Southern Asian isolates and at all amongst n=136 Eastern African isolates.

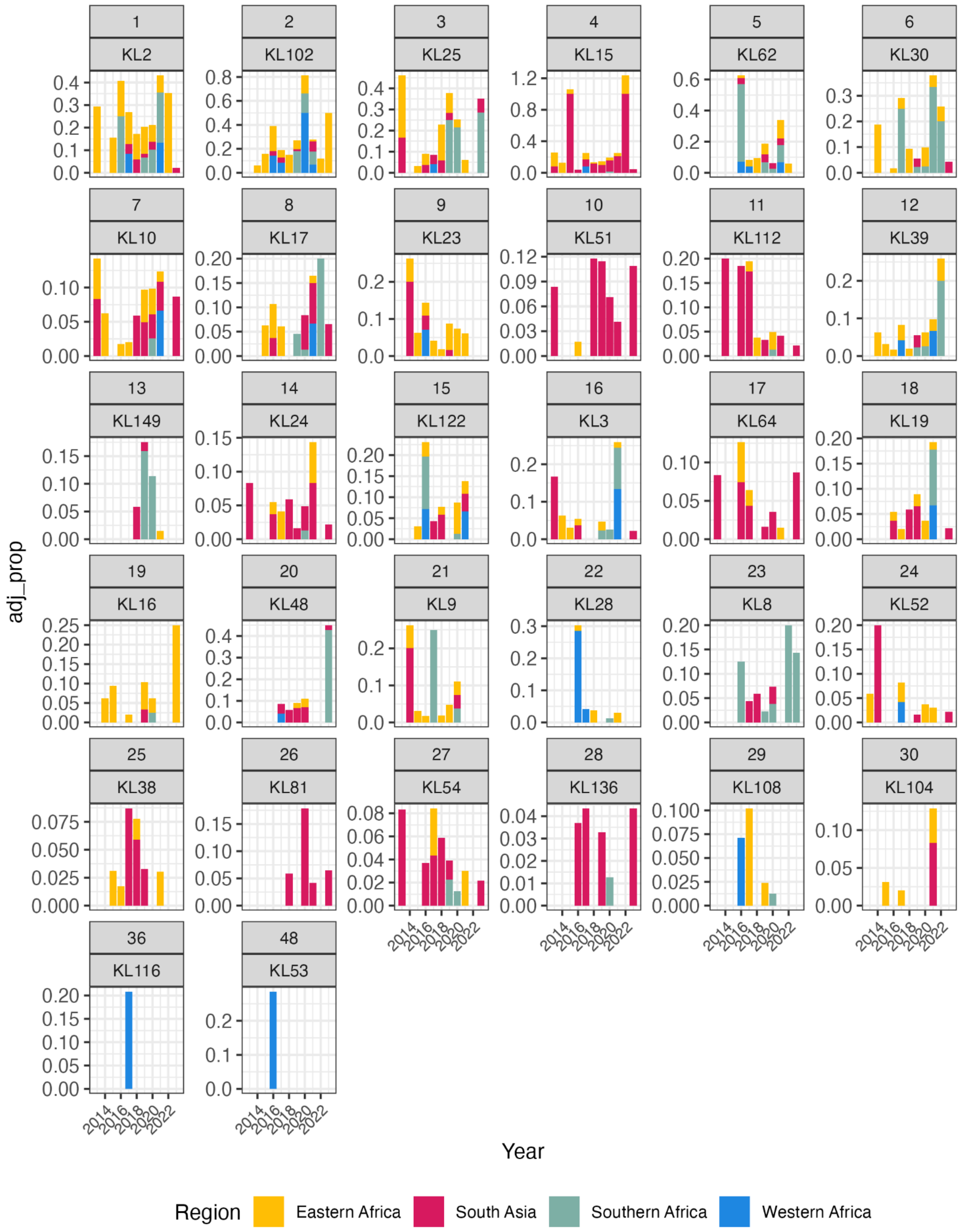

#### Appendix S1.4: Crude cluster-adjusted annual frequency per O type per region.

Bars show the crude frequency of each O type per region, using cluster-adjusted counts. Regions are coloured by inset legend. These estimates are pooled across all available studies for that region and year, i.e. the total cluster-adjusted count of isolates per O type per year in a given region, divided by the total number of unique clusters per year in that region. Plots are labelled with the O type, and the rank of that O type according to global Bayesian cluster-adjusted prevalence estimates.

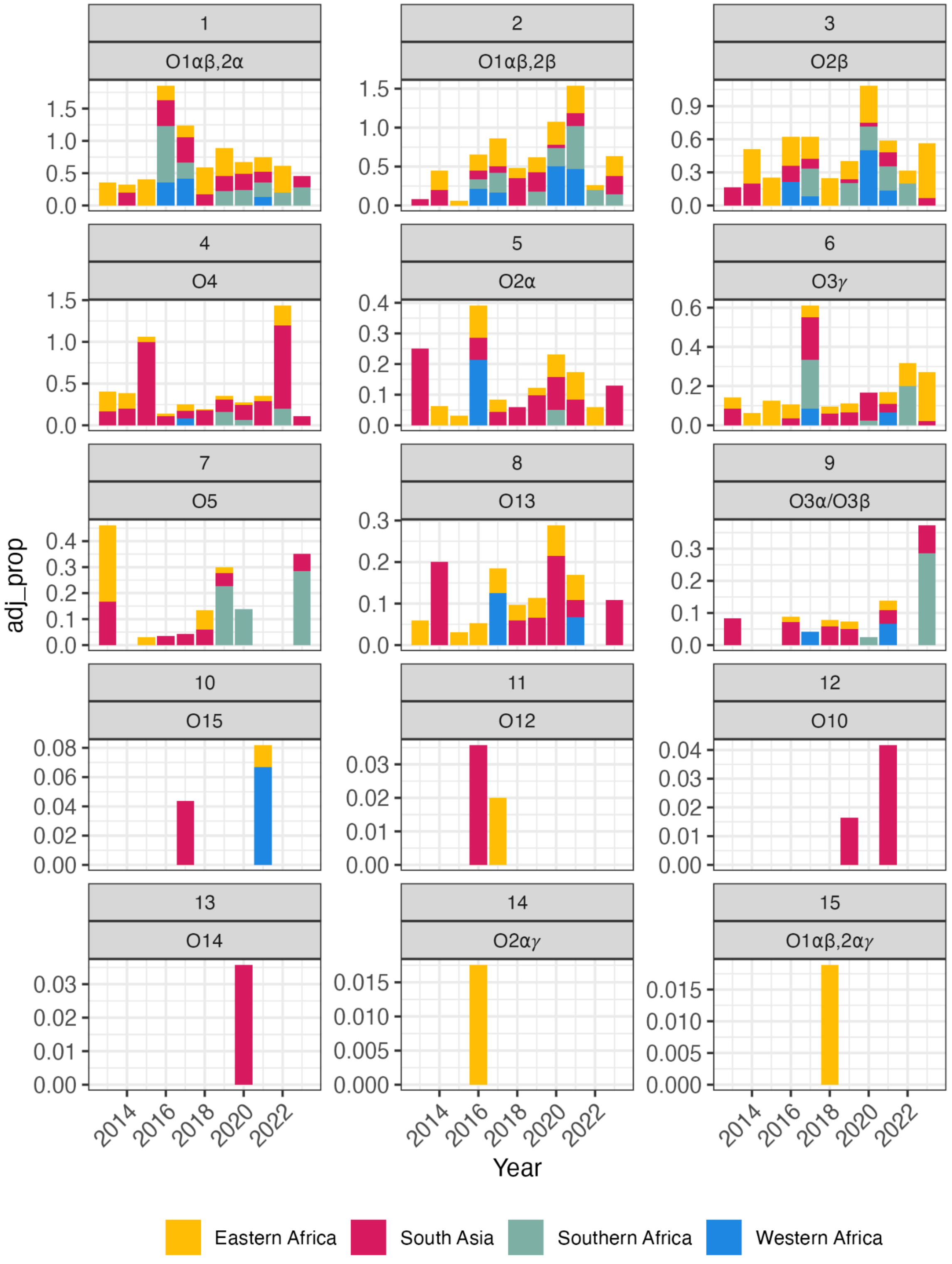

**Appendix S1.5: Region distribution of K and ST combinations associated with O types that demonstrate regional variation.**

Each plot represents the full set of isolates carrying the O type indicated in the title; each point represents a single isolate with this O type in combination with a specific sequence type (ST, indicated by the row) and K locus (KL, indicated by the column).

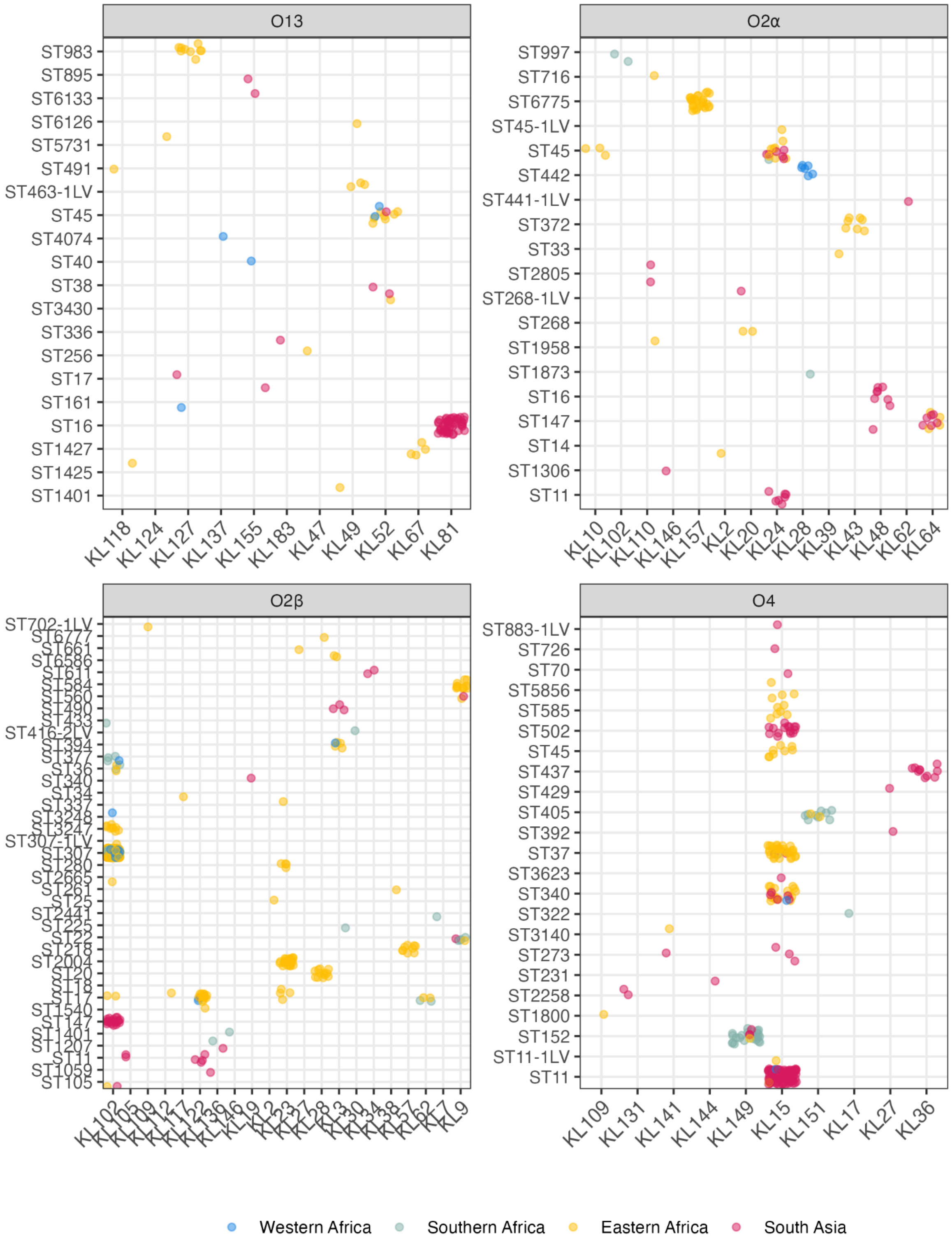

#### Appendix S1.6: Definition of genetically inferred O types used in the analysis

The unit of analysis for predicted O antigen data in this study was predicted O type, defined from the Kaptive v fields ‘O_locus’ and ‘O_type’ as shown in this table. Due to small numbers we grouped O1α,2α and O1α,2β with their matching O1αβ subtypes, i.e. O1αβ,2α and O1αβ,2β as shown in the ‘Model group’ column.

| Model group | O subtype | Kaptive v3.0 field: O_locus | Kaptive v3.0 field: O_type | n |
|---|---|---|---|---|
| O1αβ,2α | O1αβ,2α | O1/O2v1 | O1ab | 499 |
| | O1α,2α | O1/O2v1 | O1a | 1 |
| O1αβ,2β | O1αβ,2β | O1/O2v2 | O1ab | 342 |
| | O1α,2β | O1/O2v2 | O1a | 1 |
| O1αβ,2αγ | O1αβ,2αγ | O1/O2v3 | O1ab | 4 |
| O2α | O2α | O1/O2v1 | O2a | 99 |
| O2β | O2β | O1/O2v2 | O2afg | 471 |
| O2αγ | O2αγ | O1/O2v3 | O2a | 1 |
| O3α/O3β | O3α/O3β | O3/O3a | O3/O3a | 35 |
| O3γ | O3γ | O3b | O3b | 54 |
| O4 | O4 | O4 | O4 | 251 |
| O5 | O5 | O5 | O5 | 68 |
| O10 | O10 | OL103 | OL103 | 7 |
| O12 | O12 | O12 | O12 | 2 |
| O13 | O13 | OL13 | O13 | 91 |
| O14 | O14 | OL102 | OL102 | 1 |
| O15 | O15 | OL104 | OL104 | 3 |

### Appendix S2. Sensitivity analyses.

#### Appendix S2.1 Sensitivity of K locus global ranks to study inclusion.

Plots show the results of a leave-one-out analysis. In the first matrix, each column shows the global (overall) ranking of K loci, based on mean prevalence estimates modelled from cluster-adjusted counts, including either all studies (column 1) or all-but-one study (other columns). Each column is labelled by the study that was excluded from the meta-analysis. Cells are coloured (as per inset legend) to illustrate the rank of each K locus (row) in each meta-analysis (column). All K loci ranked in the top-20 for any meta-analysis are included. In the second matrix, cells show the coverage of the global top-20 from the full analysis.

Sensitivity analysis

| Excluded study | None | AKU | BabyGERMS | BARNARDS | CHRF | DH | GBS-COP | Kilifi | MBIRA | MLW | NIMBIplus | NeoBAC | NeoOBS-India | SPINZ |
|---|---|---|---|---|---|---|---|---|---|---|---|---|---|---|
| KL2 | 1 | 2 | 1 | 1 | 1 | 1 | 1 | 1 | 2 | 3 | 2 | 3 | 1 | 1 |
| KL102 | 2 | 1 | 2 | 2 | 2 | 2 | 2 | 2 | 1 | 1 | 1 | 1 | 2 | 3 |
| KL25 | 3 | 3 | 3 | 3 | 3 | 3 | 3 | 3 | 3 | 2 | 3 | 2 | 3 | 2 |
| KL15 | 4 | 4 | 4 | 4 | 5 | 4 | 4 | 4 | 4 | 4 | 4 | 4 | 4 | 4 |
| KL62 | 5 | 5 | 5 | 5 | 4 | 5 | 5 | 5 | 5 | 5 | 5 | 5 | 5 | 5 |
| KL30 | 6 | 6 | 6 | 6 | 6 | 6 | 6 | 6 | 6 | 12 | 6 | 6 | 6 | 6 |
| KL10 | 7 | 10 | 10 | 8 | 9 | 8 | 8 | 8 | 7 | 8 | 7 | 8 | 8 | 7 |
| KL17 | 8 | 8 | 8 | 7 | 7 | 7 | 7 | 7 | 11 | 6 | 8 | 7 | 7 | 9 |
| KL23 | 9 | 7 | 7 | 9 | 8 | 9 | 9 | 12 | 10 | 10 | 9 | 10 | 9 | 10 |
| KL51 | 10 | 19 | 11 | 11 | | 12 | 12 | 11 | 8 | 11 | 14 | 11 | 13 | 12 |
| KL112 | 11 | 9 | 9 | | 10 | 10 | 10 | 9 | 9 | 7 | 10 | 9 | 10 | 8 |
| KL39 | 12 | 12 | 16 | 13 | 11 | 11 | 13 | 18 | 12 | 15 | 15 | 14 | 12 | 15 |
| KL149 | 13 | 11 | | 12 | 12 | 13 | | 10 | 13 | 9 | 13 | 13 | 11 | 13 |
| KL24 | 14 | 16 | 15 | 19 | 16 | 20 | 16 | 14 | 18 | 17 | 17 | 17 | 17 | 20 |
| KL122 | 15 | 13 | 14 | 15 | 13 | 15 | 11 | 17 | 17 | 13 | 11 | 15 | 15 | 14 |
| KL3 | 16 | 14 | 13 | 10 | 15 | 16 | 15 | 16 | 20 | 14 | 12 | 12 | 14 | 11 |
| KL64 | 17 | | 12 | | 18 | 14 | 14 | 13 | 14 | 16 | 16 | 16 | 16 | 17 |
| KL19 | 18 | | 18 | 16 | 20 | 17 | 20 | 20 | | | 20 | | | |
| KL16 | 19 | 17 | 19 | 14 | 14 | 18 | 19 | | 16 | | 19 | 20 | 20 | 16 |
| KL48 | 20 | 18 | 17 | 17 | | 19 | 17 | 15 | 15 | 18 | | 19 | 19 | 19 |
| KL9 | | 15 | | 18 | 17 | | 18 | | 19 | | 18 | 18 | 18 | 18 |
| KL28 | | | | | 19 | | | | | | | | | |
| KL8 | | 20 | | | | | | | | 19 | | | | |
| KL81 | | | 20 | 20 | | | | 19 | | | | | | |
| KL54 | | | | | | | | | | 20 | | | | |

| Excluded study | None | AKU | BabyGERMS | BARNARDS | CHRF | DH | GBS-COP | Kilifi | MBIRA | MLW | NIMBIplus | NeoBAC | NeoOBS-India | SPINZ |
|---|---|---|---|---|---|---|---|---|---|---|---|---|---|---|
| Global | 72.77 | 73.58 | 72.38 | 75.88 | 73.78 | 72.88 | 72.37 | 72.76 | 74.96 | 70.16 | 73.61 | 73.63 | 72.53 | 69.06 |
| South Asia | 67.79 | 67.75 | 66.01 | 66.76 | 60.19 | 67.89 | 65.97 | 66.11 | 67.7 | 65.99 | 66.07 | 66.08 | 66.1 | 65.99 |
| Eastern Africa | 75.68 | 76.54 | 76.51 | 79.05 | 76.59 | 75.66 | 76.5 | 76.72 | 80.42 | 72.6 | 76.51 | 77.84 | 75.64 | 70.28 |
| Southern Africa | 76.41 | 76.62 | 71.74 | 77.12 | 76.53 | 76.44 | 72.16 | 76.59 | 75.55 | 76.55 | 82.25 | 76.53 | 76.55 | 76.45 |
| Western Africa | 50.49 | 49.84 | 49.8 | 78.95 | 49.36 | 50.55 | 49.74 | 49.85 | 35.47 | 49.7 | 49.88 | 50.01 | 50 | 49.69 |

Excluded study

**Appendix S2.2 Sensitivity of O type global ranks to study inclusion.**

Plots show the results of a leave-one-out analysis. Top matrix shows the results of a leave-one-out analysis. Each column shows the global (overall) ranking of O types, based on mean prevalence estimates modelled from cluster-adjusted counts, including either all studies (column 1) or all-but-one study (other columns). Each column is labelled by the study that was excluded from the meta-analysis. Cells are coloured (as per inset legend) to illustrate the rank of each O type (row) in each meta-analysis (column). Only the top nine O types are shown (the other six loci are excluded as they were rarely observed: O1αβ,2αγ, n=4; O2αγ, n=1; O12, n=2; O10, n=7; O14, n=1; O15, n=2). In the matrices below, cells show the coverage of the global top-4 or top-5 from the full analysis.

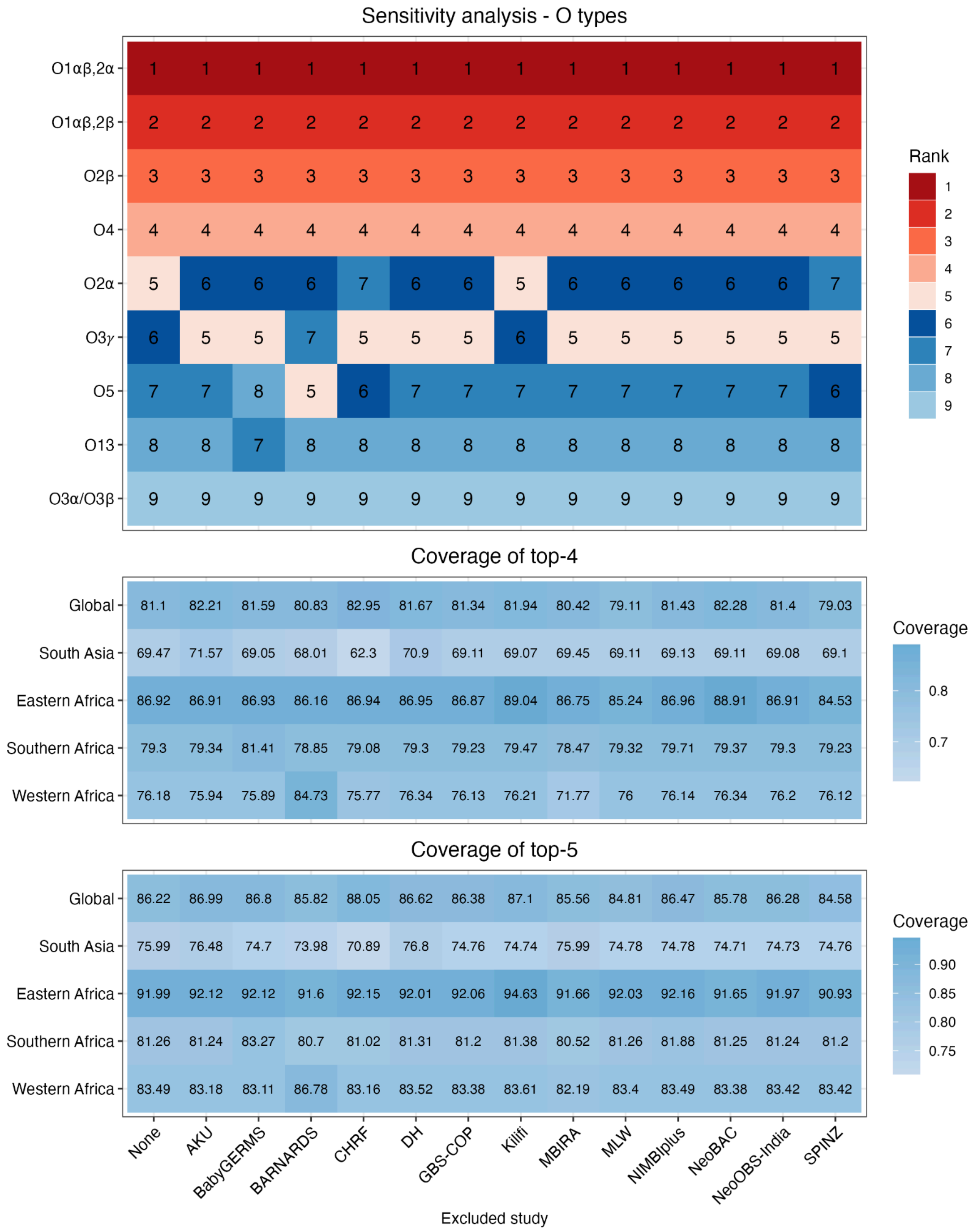

#### Appendix S2.3 Effect of cluster adjustment on crude proportions per site.

Plots show crude proportion per site, estimated using raw (x-axis) vs cluster-adjusted (y-axis) counts, for the 4 most common K loci **(a-d)** and O types **(e-h)**. The impact of cluster-adjustment on the global prevalence estimates are shown in **Appendix S2.4**.

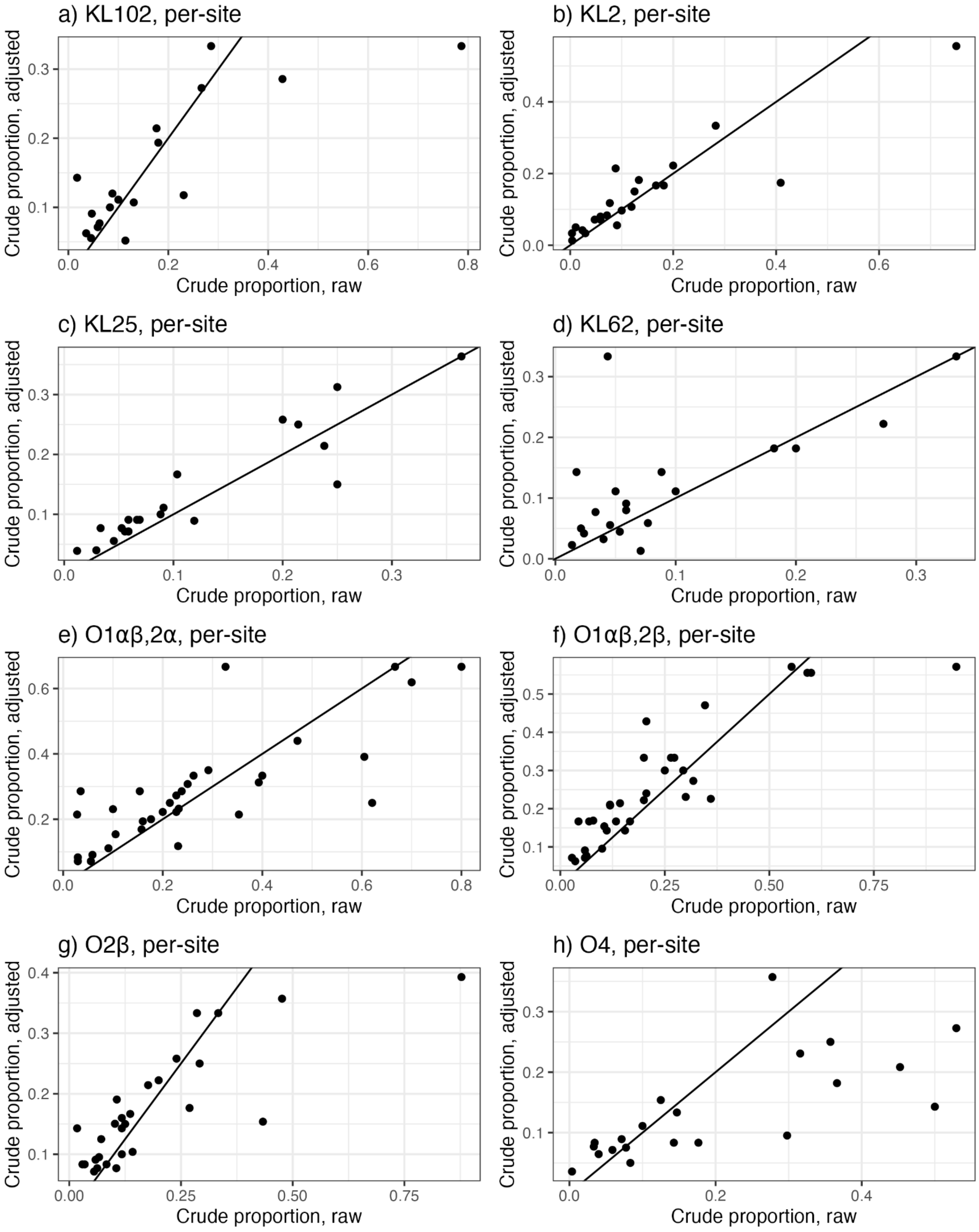

#### Appendix S2.4 Effect of cluster adjustment on Bayesian modelled estimates of global K locus prevalence.

Global (overall) prevalence estimates modelled for **(a)** each K locus or **(b)** each predicted O type, using either raw counts (x-axis) vs cluster-adjusted counts (y-axis).

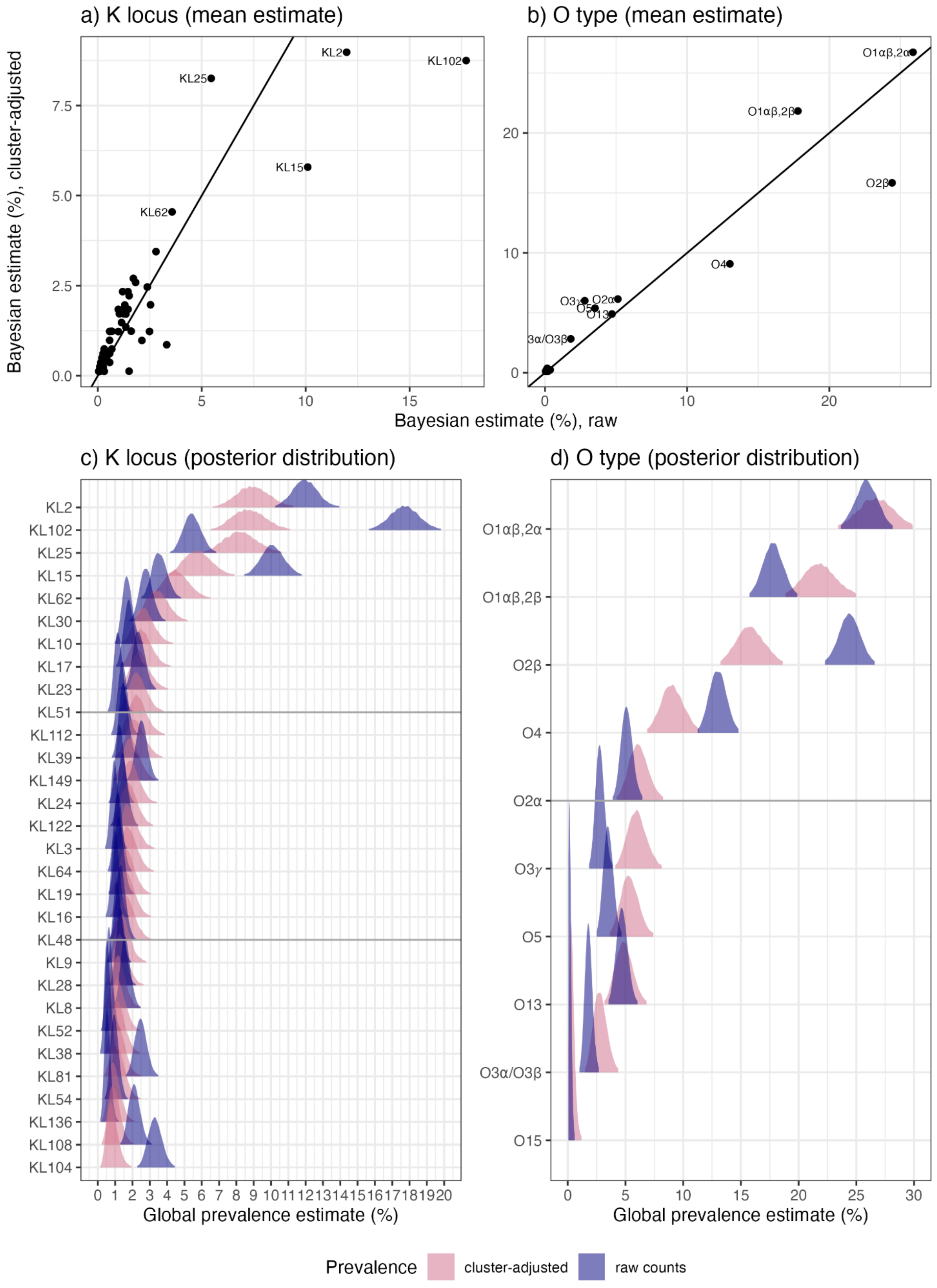

#### Appendix S2.5 Sensitivity analysis showing effect of temporal clustering thresholds on K locus proportions and ranks (modelled global estimates).

The clustering threshold used for all primary analyses was 28 days; modelling was repeated using the 365-day threshold to assess sensitivity to clustering parameters. Data plotted here summarise the Bayesian modelled estimates for global prevalence, using counts adjusted for clustering using different temporal thresholds (≤28 days vs ≤365 days). In panels (**a-d**) each point represents a single K locus or O type. Panels (**e-f**) show the posterior distributions of prevalence estimates.

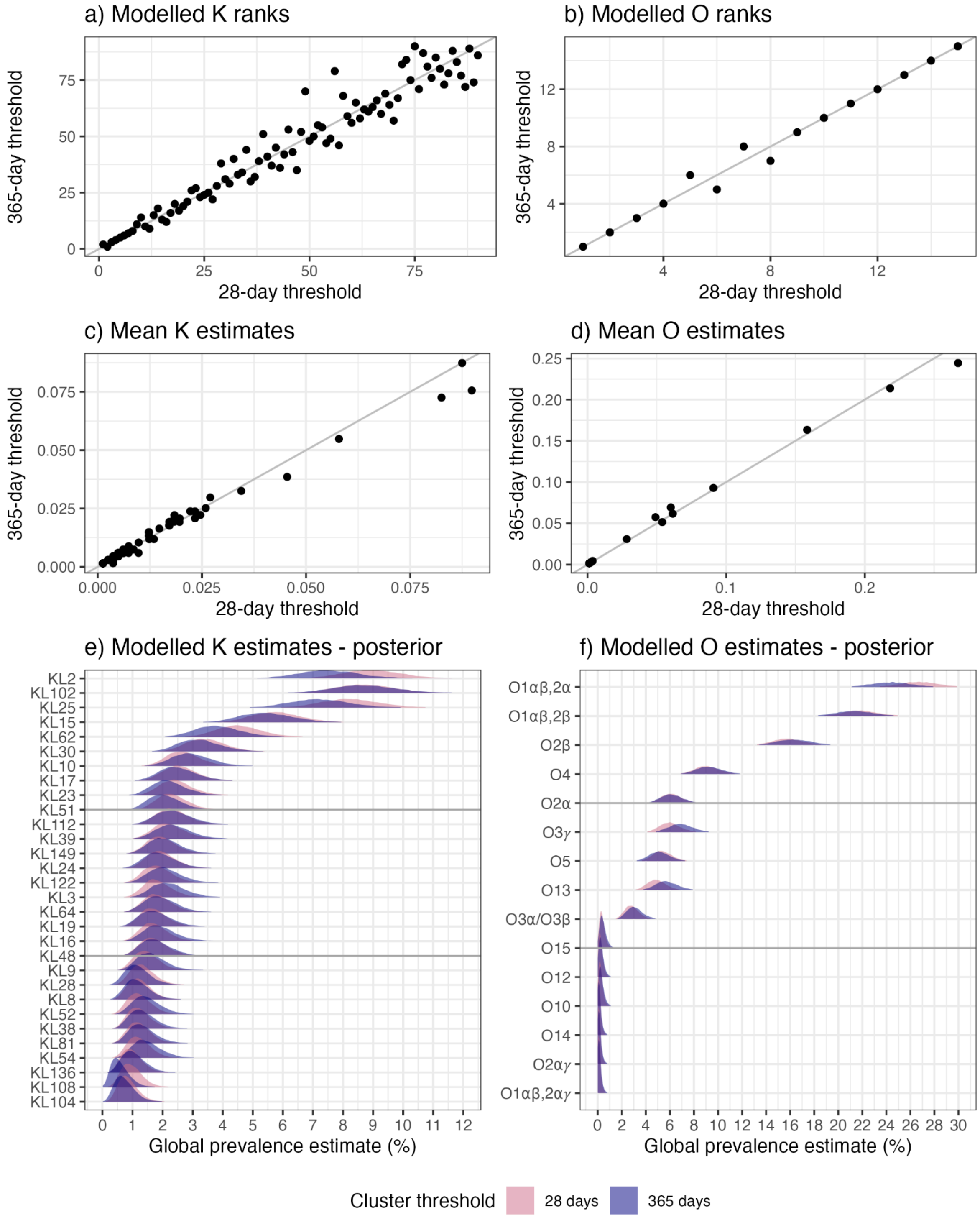

#### Appendix S2.6 Sensitivity analysis showing effect of temporal clustering thresholds on K locus proportions and ranks (crude proportions).

Plotted data are based on crude proportions (rather than Bayesian modelled estimates of prevalence), using a SNP threshold of 10. The thresholds used for primary analysis were 28 days (red) and 10 SNPs.

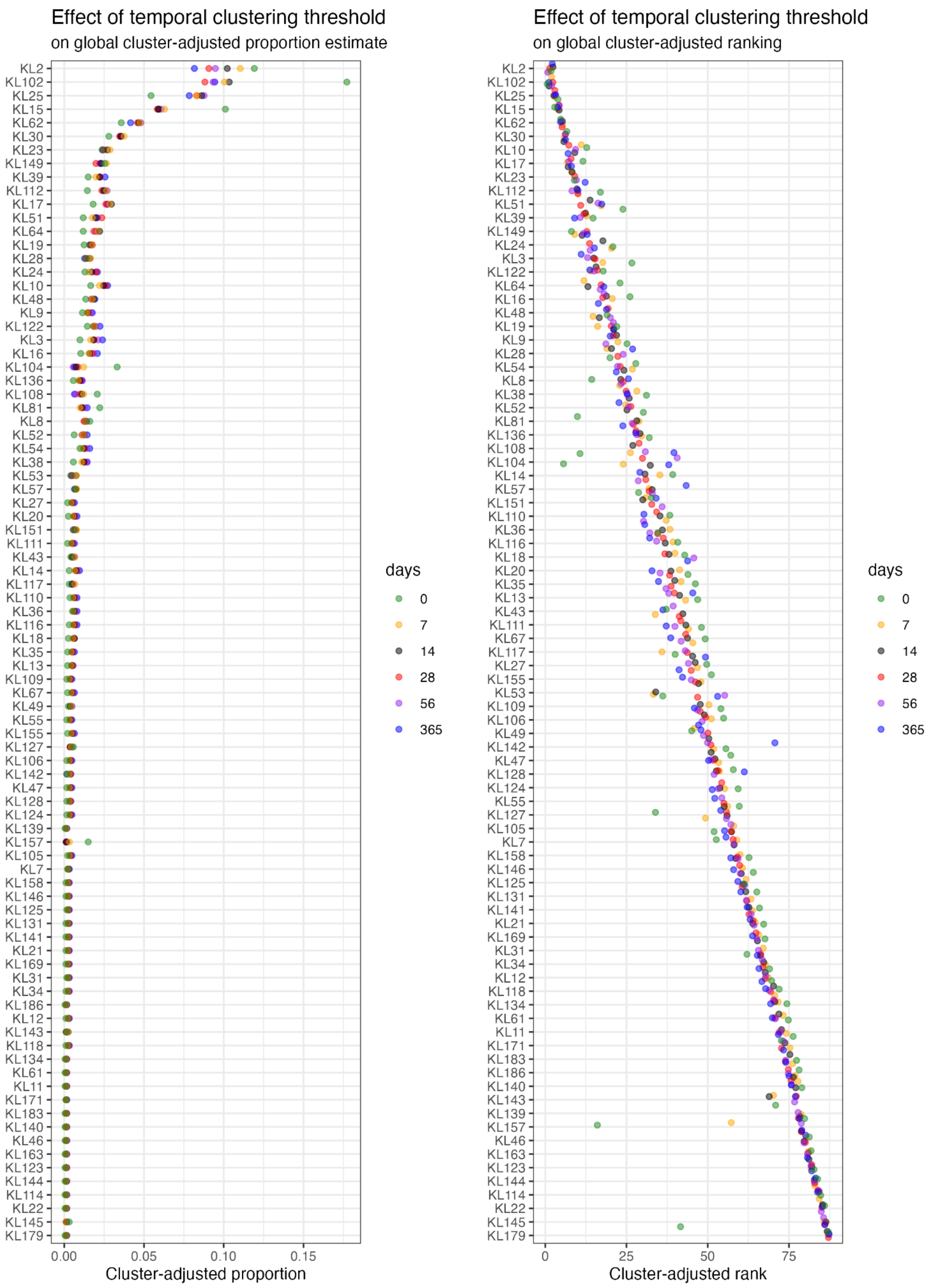

#### Appendix S2.7 Sensitivity analysis showing effect of SNP clustering thresholds on K locus proportions and ranks (crude proportions).

Plotted data are based on crude proportions (rather than Bayesian modelled estimates of prevalence), using a temporal threshold of 4 weeks. The thresholds used for primary analysis were 10 SNPs (red) and 4 weeks.

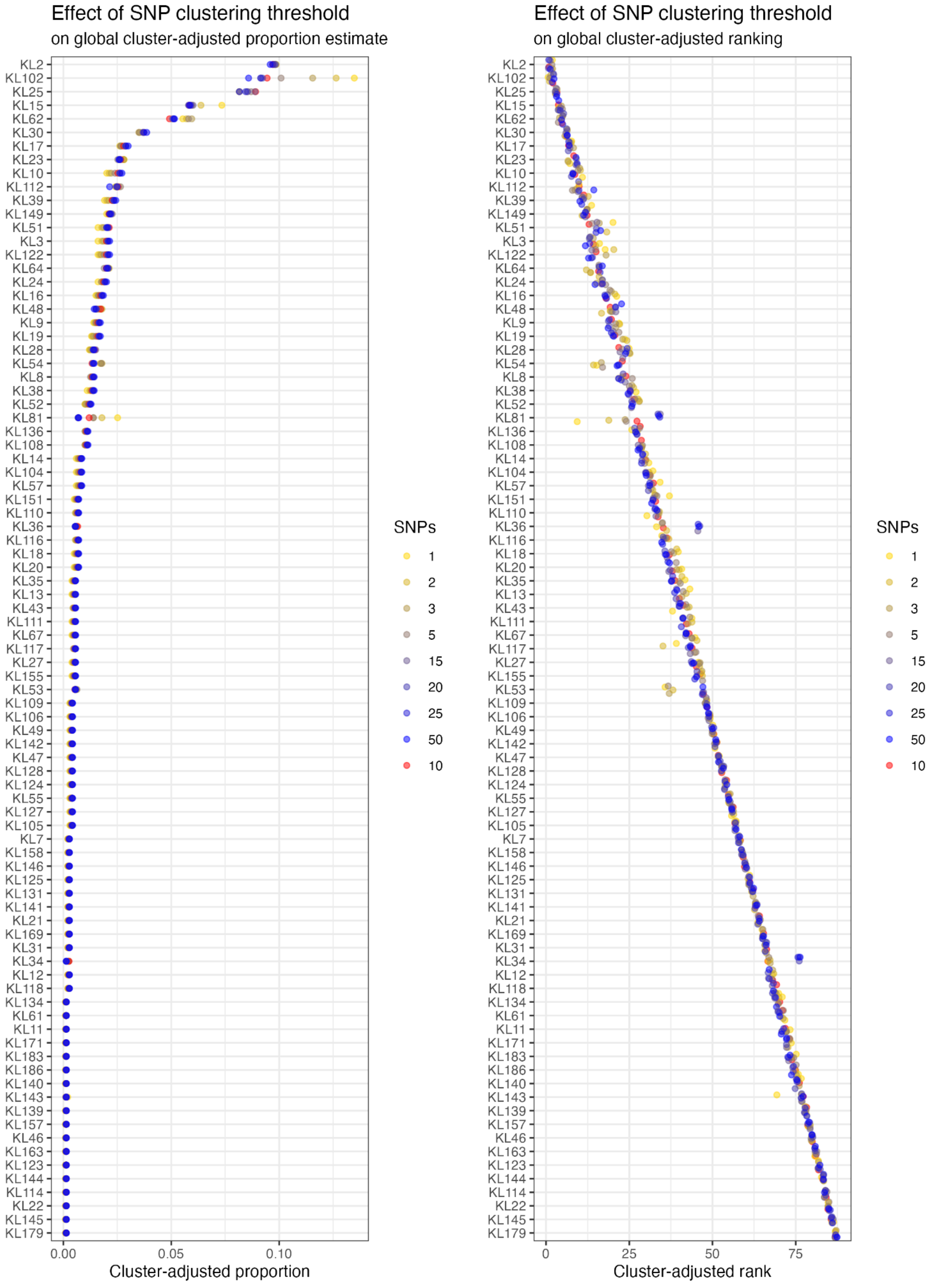

#### Appendix S2.8: Coverage estimates for KL set 1 (global top 20)

Cumulative coverage estimates for the top 20 K loci (KL), ordered by cluster-adjusted global prevalence estimate. The first panel shows estimates for all cases (“All”), remaining panels show coverage estimated for the subset of cases indicated in the panel title (ESBL, extended-spectrum beta-lactamase; CP, carbapenemase-producing). Coverage estimated from raw counts are shown as solid lines, and from cluster-adjusted counts as dashed lines, each with 95% credible intervals shaded.

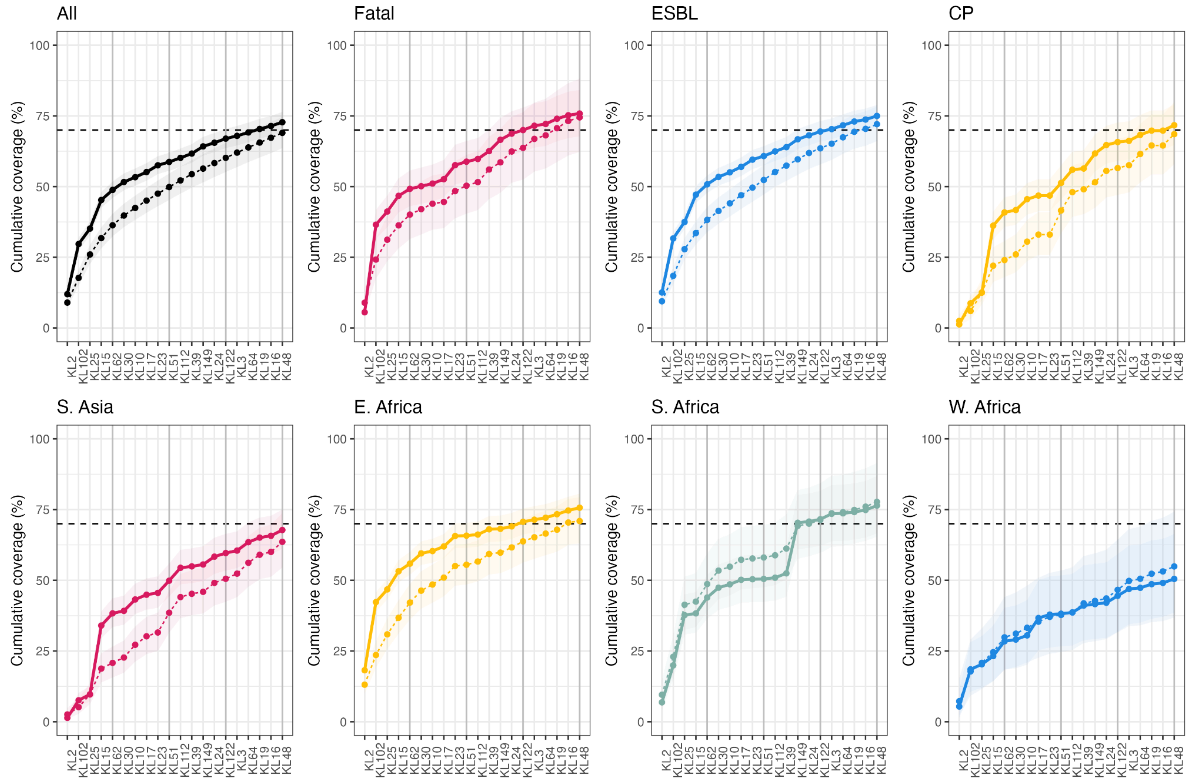

#### Appendix S2.9: Coverage estimates for KL set 2 (top-8 per region)

Cumulative coverage estimates for KL set 2, comprising the top-8 K loci within each region, ranked according to cluster-adjusted global prevalence estimate. The first panel shows estimates for all cases ("All"), remaining panels show coverage estimated for the subset of cases indicated in the panel title (ESBL, extended-spectrum beta-lactamase; CP, carbapenemase-producing). Coverage estimated from raw counts are shown as solid lines, and from cluster-adjusted counts as dashed lines, each with 95% credible intervals shaded.

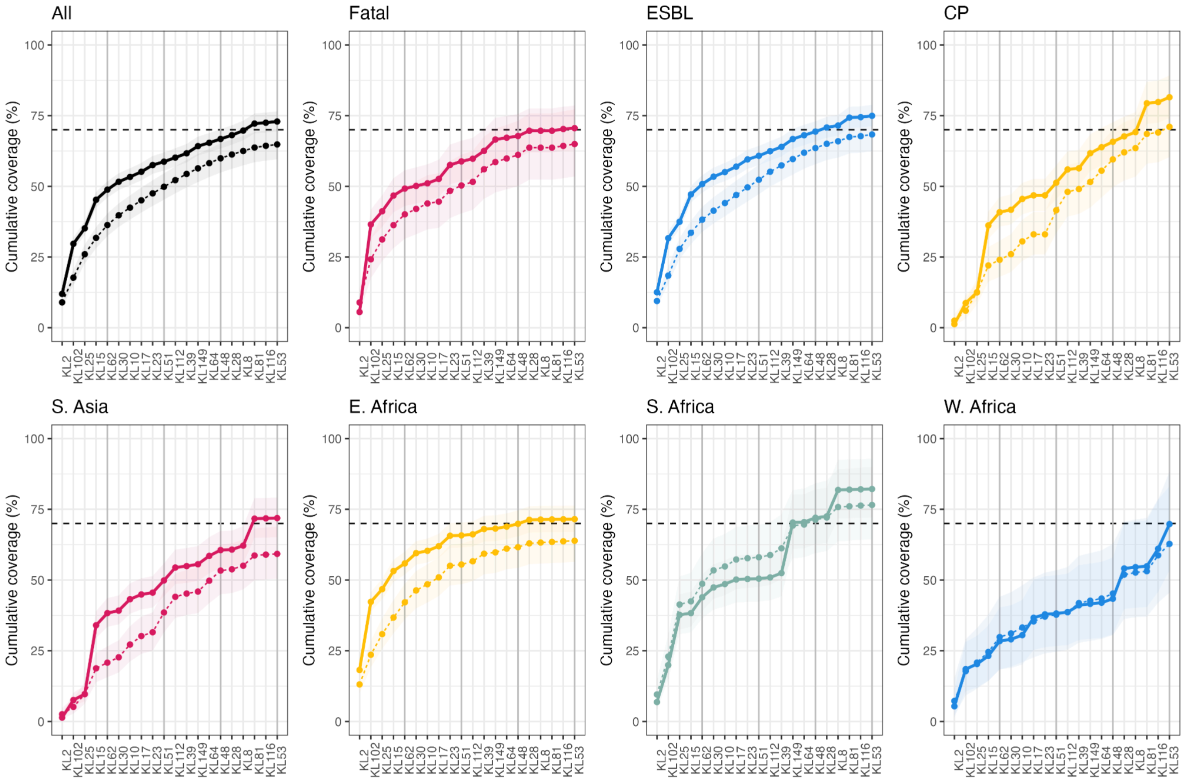

#### Appendix S2.10: Coverage estimates for O types

Cumulative coverage estimates for the top 10 O types, ordered by cluster-adjusted global prevalence estimate. The first panel shows estimates for all cases (“All”), remaining panels show coverage estimated for the subset of cases indicated in the panel title (ESBL, extended-spectrum beta-lactamase; CP, carbapenemase-producing). Coverage estimated from raw counts are shown as solid lines, and from cluster-adjusted counts as dashed lines, each with 95% credible intervals shaded.

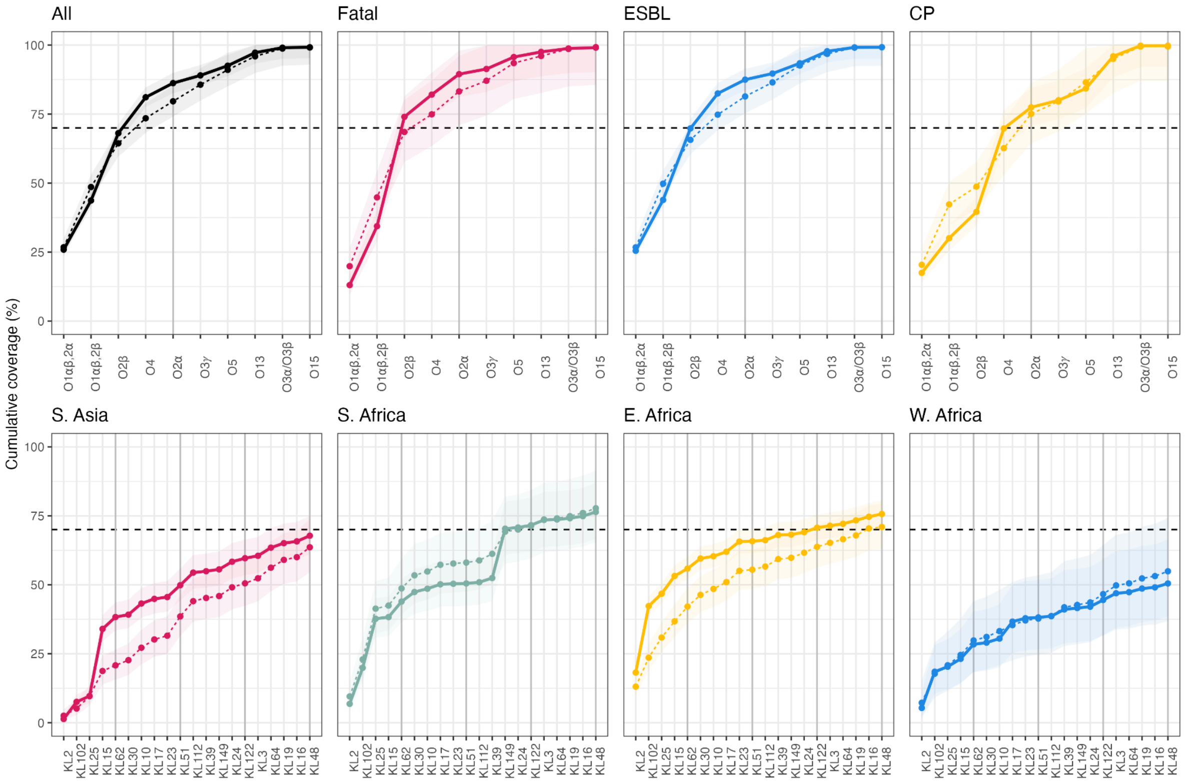

#### Appendix S3. Model Diagnostics.

We evaluated three key diagnostic metrics to ensure the robustness of our Bayesian inference: Rhat, the effective sample size ratio, and divergent transitions.

Rhat is a metric of convergence. It compares the between-chain variance to the within-chain variance, providing an indication of whether multiple Markov chains have converged to the same target distribution. Values close to 1 suggest that the chains have mixed well; values substantially above 1 may indicate potential issues with convergence. In our models, we checked that the maximum Rhat were under 1.01. All values were markedly below this (see table below).

The effective sample size ratio measures sampling efficiency by estimating the number of effectively independent samples obtained from the correlated MCMC draws. Since autocorrelation reduces the number of independent samples, a higher ratio indicates that the sampling process is efficient and that the posterior estimates are reliable. We computed the minimum effective sample size ratio for each model to confirm that our inference is based on a sufficiently large number of independent draws. All models, with the exception of models a and b, had minimum effective sample size ratios of 0.1 (see table below) or above indicating samples were sufficiently independent. These models were rerun with more iterations as described in the Methods. When re-run, those models which still had a ratio of less than 0.1 were then inspected closely to determine whether this was a feature of all effective sample size ratios, or rare aberrations. It was determined that these low ratios impacted very few (as low as 1) model parameters, and as such the models were deemed acceptable.

Divergent transitions occur when the sampler encounters regions in the posterior distribution with challenging geometry, which can lead to biased estimates. The absence of any divergent transitions in our diagnostics confirms that the sampler was able to explore the posterior landscape smoothly and without numerical instabilities.

Model diagnostics for all Bayesian models

**Primary models**

| Model ID | subgroup | counts | days | antigen | max Rhat | min Neff_ratio |
|---|---|---|---|---|---|---|
| a | CP | adj | 28 | K | 1.00088238 | 0.03796967 |
| b | CP | raw | 28 | K | 1.00140556 | 0.0548587 |
| c | CP | adj | 28 | O | 1.00202916 | 0.1828553 |
| d | CP | raw | 28 | O | 1.00121229 | 0.18043056 |
| e | ESBL | adj | 28 | K | 1.00180625 | 0.23567658 |
| f | ESBL | raw | 28 | K | 1.00178756 | 0.25413868 |
| g | ESBL | adj | 28 | O | 1.00171688 | 0.23119961 |
| h | ESBL | raw | 28 | O | 1.00202841 | 0.26851334 |
| i | Fatal | adj | 28 | K | 1.00203937 | 0.19809457 |
| j | Fatal | raw | 28 | K | 1.00277878 | 0.1854011 |
| k | Fatal | adj | 28 | O | 1.001967 | 0.2824846 |
| l | Fatal | raw | 28 | O | 1.00137218 | 0.24379735 |
| m | Full | adj | 28 | K | 1.00269449 | 0.2116118 |
| n | Full | raw | 28 | K | 1.00216508 | 0.24603945 |
| o | Full | adj | 28 | O | 1.00151937 | 0.29722619 |
| p | Full | raw | 28 | O | 1.00196789 | 0.27290005 |
| q | Full | adj | 365 | K | 1.00146703 | 0.246428 |
| r | Full | raw | 365 | K | 1.00189497 | 0.253698 |
| s | Full | adj | 365 | O | 1.00166388 | 0.29827219 |
| t | Full | raw | 365 | O | 1.00144951 | 0.26270787 |

**Leave-one-out analysis**

| Model ID | study left out | type | days | antigen | max Rhat | min Neff_ratio |
|---|---|---|---|---|---|---|
| 1 | DH | adj | 28 | K | 1.00201008 | 0.22653089 |
| 2 | DH | raw | 28 | K | 1.00162068 | 0.25393721 |
| 3 | AKU | adj | 28 | K | 1.00193839 | 0.22544558 |
| 4 | AKU | raw | 28 | K | 1.00244137 | 0.24700401 |
| 5 | Baby GERMS-SA | adj | 28 | K | 1.00124264 | 0.23064113 |
| 6 | Baby GERMS-SA | raw | 28 | K | 1.00255461 | 0.24534982 |
| 7 | BARNARDS | adj | 28 | K | 1.00167169 | 0.25459179 |
| 8 | BARNARDS | raw | 28 | K | 1.00177469 | 0.23131859 |
| 9 | NIMBIplus | adj | 28 | K | 1.00194693 | 0.18831693 |
| 10 | NIMBIplus | raw | 28 | K | 1.00154699 | 0.23196231 |

| | | | | | | |
|---|---|---|---|---|---|---|
| 11 | CHRF | adj | 28 | K | 1.00221728 | 0.21047287 |
| 12 | CHRF | raw | 28 | K | 1.00224478 | 0.25348302 |
| 13 | GBS-COP | adj | 28 | K | 1.00338847 | 0.20016976 |
| 14 | GBS-COP | raw | 28 | K | 1.00167867 | 0.25835135 |
| 15 | Kilifi | adj | 28 | K | 1.00275235 | 0.20073946 |
| 16 | Kilifi | raw | 28 | K | 1.0017595 | 0.26507194 |
| 17 | MBIRA | adj | 28 | K | 1.00463533 | 0.16901181 |
| 18 | MBIRA | raw | 28 | K | 1.00304928 | 0.2466877 |
| 19 | MLW | adj | 28 | K | 1.00144484 | 0.21951426 |
| 20 | MLW | raw | 28 | K | 1.0015223 | 0.25765252 |
| 21 | NeoBAC | adj | 28 | K | 1.00188468 | 0.21835634 |
| 22 | NeoBAC | raw | 28 | K | 1.00205798 | 0.22931353 |
| 23 | SPINZ | adj | 28 | K | 1.00213139 | 0.2262595 |
| 24 | SPINZ | raw | 28 | K | 1.00169702 | 0.24546798 |
| 25 | DH | adj | 28 | O | 1.00203495 | 0.27384274 |
| 26 | DH | raw | 28 | O | 1.00106341 | 0.25978561 |
| 27 | AKU | adj | 28 | O | 1.00200069 | 0.26262671 |
| 28 | AKU | raw | 28 | O | 1.00148952 | 0.26859157 |
| 29 | Baby GERMS-SA | adj | 28 | O | 1.00128027 | 0.27731336 |
| 30 | Baby GERMS-SA | raw | 28 | O | 1.00166639 | 0.27880282 |
| 31 | BARNARDS | adj | 28 | O | 1.00236419 | 0.27353777 |
| 32 | BARNARDS | raw | 28 | O | 1.00178975 | 0.27661917 |
| 33 | NIMBIplus | adj | 28 | O | 1.00095957 | 0.28238137 |
| 34 | NIMBIplus | raw | 28 | O | 1.00121788 | 0.26769635 |
| 35 | CHRF | adj | 28 | O | 1.00135568 | 0.27264555 |
| 36 | CHRF | raw | 28 | O | 1.0015541 | 0.25169272 |
| 37 | GBS-COP | adj | 28 | O | 1.00139764 | 0.30402441 |
| 38 | GBS-COP | raw | 28 | O | 1.00199053 | 0.27497652 |
| 39 | Kilifi | adj | 28 | O | 1.00176647 | 0.26223239 |
| 40 | Kilifi | raw | 28 | O | 1.00099135 | 0.29196706 |
| 41 | MBIRA | adj | 28 | O | 1.00202812 | 0.29837529 |
| 42 | MBIRA | raw | 28 | O | 1.00199366 | 0.25439415 |
| 43 | MLW | adj | 28 | O | 1.00106562 | 0.28017804 |
| 44 | MLW | raw | 28 | O | 1.00133002 | 0.26457843 |
| 45 | NeoBAC | adj | 28 | O | 1.00119297 | 0.27130553 |
| 46 | NeoBAC | raw | 28 | O | 1.00144345 | 0.27021013 |
| 47 | SPINZ | adj | 28 | O | 1.00132456 | 0.28506661 |
| 48 | SPINZ | raw | 28 | O | 1.00175768 | 0.25543517 |
